# A Nine-Year Analysis of WHO Critical Priority Pathogens from the Tunisian AMR Surveillance System

**DOI:** 10.64898/2026.03.23.26349077

**Authors:** Dana Itani, Laura T Phillips, Sara Kotb Tolba, Wafaa Achour, Hanen Smaoui, Lamia Thabet, Meriam Zribi, Ebenezer Foster-Nyarko, Kathryn E Holt, Ilhem Boutiba – Ben Boubaker

**Author notes:** **Corresponding author contact: Dana Itani**, Department of Infection Biology, Faculty of Infectious and Tropical Diseases, London School of Hygiene and Tropical Medicine, London, UK. These authors contributed equally to this work (Wafaa Achour, Hanen Smaoui, Lamia Thabet, Meriam Zribi).

## Abstract

**Background:** Antimicrobial resistance (AMR) surveillance is essential for quantifying and monitoring the burden of AMR among World Health Organization (WHO) priority pathogens. We analysed Tunisian AMR surveillance system (TARSS) data across five sentinel hospitals from 2014 to 2022.

**Methods:** We conducted a retrospective isolate-level analysis for *Escherichia coli*, *Klebsiella pneumoniae*, *Pseudomonas aeruginosa*, and *Acinetobacter* spp. Temporal, ward, and specimen associations were quantified using multivariable logistic regression models. Sex and age categories were explored in secondary models due to missingness. Temporal trends were assessed using Cochran–Armitage test, and co-resistance was summarised for third-generation cephalosporin and carbapenem phenotypes. We also evaluated temporal dynamics of 3GCR and CR profiles.

**Results:** A total of 35,525 *E. coli*, 14,325 *K. pneumoniae*, 9,679 *P. aeruginosa,* and 5,597 *Acinetobacter* spp. were reported to TARSS between 2014 and 2022. Mean annual MDR prevalence was high for *Acinetobacter* spp. (85.1%), moderate for *K. pneumoniae* (45.5%) and for *P. aeruginosa* (27.1%), and lower for *E. coli* (17.5%). Adjusted models indicated increased odds of resistance to several antibiotics, whereas *E. coli* showed decreased odds. Intensive care unit (ICU) and blood isolates were associated with higher odds of resistance in all pathogens.

**Conclusion:** This nine-year multi-hospital analysis reveals a high prevalence of AMR across the four WHO priority pathogens, settings, and specimen types, with increasing resistance for some pathogen-antibiotic combinations. The higher odds of clinically important resistance amongst ICU and blood isolates support the use of ward-level antibiograms and stratified stewardship and infection prevention measures.

## Introduction

Antimicrobial resistance (AMR) is a global public health challenge with the potential to result in up to ten million fatalities annually by 2050 (1). Surveillance is central to estimating the burden of resistance, guiding empirical treatment, monitoring trends, detecting new resistance patterns, and evaluating interventions. Surveillance is recognised as a cornerstone of action against AMR, as emphasised by the World Health Organization (WHO) Global Action Plan on AMR (2–4). However, in low- and middle-income countries (LMICs), the lack of quality-assured data and weak surveillance infrastructure obscure the true burden of AMR and impede action (5).

Tunisia, a country in northern Africa classified by the World Bank as lower-middle income, has historically lacked AMR data, leaving critical gaps in understanding resistance patterns for bacterial pathogens (6). To address this, the Tunisian Ministry of Health launched the National Action Plan on AMR and established the Tunisian AMR Surveillance System (TARSS) in 2019 (7). This system is based on the WHO Global Antimicrobial Surveillance System (GLASS) model and comprises a National Collaborating Centre (NCC), National Reference Laboratory (NRL), and surveillance sites (SS) (3). TARSS involves passive routine surveillance across 11 hospitals in different governorates in Tunisia, by collecting, analysing, and reporting standardised demographic, epidemiological, and microbiological data for priority pathogens. Priority pathogens include *Acinetobacter* spp., *Pseudomonas aeruginosa,* and the Enterobacterales *Klebsiella pneumoniae* and *Escherichia coli*. These four pathogens were classified by WHO as critical priority due to their multidrug-resistant profiles and the need for new antibiotics (8).

In Tunisian hospitals, *Acinetobacter* spp., *P. aeruginosa,* and *K. pneumoniae* and *E. coli* are of major concern due to the risk of healthcare-associated infections, driving prolonged hospital stays and increased risk of mortality and morbidity, particularly among immunocompromised patients (9). In addition, there are systematic challenges related to broad-spectrum antibiotic use, infection prevention and control measures, and diagnostic delays due to limited laboratory capacity (10). Here, we present a comprehensive analysis of the AMR epidemiology of *Acinetobacter* spp., *P. aeruginosa, K. pneumoniae*, and *E. coli* across five TARSS sites from 2014 to 2022. This study aims to inform surveillance strategies and targeted interventions to mitigate AMR.

## Methods

### Ethical consideration

This study adheres to all relevant national and international ethical regulations. Ethics approval was granted by the Ministry of Health in Tunisia (Authorization No. 0000021-080000-20-2023), the internationally recognized Ethics Committee of Charles Nicolle University Hospital (FWA00032748 and IORG0011243), and the LSHTM Observational/Interventions Research Ethics Committee (Ref: 26901). The Charles Nicolle University Hospital Ethics Committee provided a waiver of individual informed consent as the study involves secondary analysis of anonymized, pre-existing surveillance data.

### Study design and setting

We conducted a retrospective study using passive surveillance data collected through TARSS from January 2014 to December 2022. The analysis included isolate-level microbiological and epidemiological data derived from clinical specimens collected from patients at five sentinel hospitals. Since TARSS was not formally established until 2019, data from 2014 to 2018 were validated by the NRL of AMR to ensure compatibility with the new TARSS reporting standards. This study was conducted and reported according to Strengthening the Reporting of Observational studies in Epidemiology (STROBE) guidelines (11).

### Surveillance network and data sources

TARSS monitors resistance in WHO GLASS priority pathogens, including *E. coli, K. pneumoniae, Acinetobacter* spp*., Staphylococcus aureus, Streptococcus pneumoniae, Salmonella* spp. and *Shigella* spp. These pathogens are isolated from multiple specimen types (blood, urine, stool, wounds, soft tissues, cerebrospinal fluid [CSF], respiratory tracts, and genital tract). Data on bacterial identification and antimicrobial susceptibility tests (AST) were electronically shared from 11 sentinel laboratories to the NRL on an annual basis. The laboratories use two data management systems: Santé Lab, a laboratory information system, and SIRscan, an automated AST reader. These systems were linked following the establishment of TARSS in 2019 to enable standardized national reporting.

Five TARSS sites participated in this study (summarized in **Supplementary Table S1**). They are located within the governorates of Tunis and Ben Arous, selected to represent diverse tertiary and specialty settings within Tunis (**Figure 1**). These sites include two general tertiary care hospitals, Charles Nicolle University Hospital, La Rabta University Hospital, as well as three specialized facilities, Centre National de Greffe de la Moelle Osseuse [CNGMO], Trauma Centre and Burns [TCB], and Children’s Hospital Bechir Hamza [CHBH]. These hospitals serve 24.5% of Tunisia’s urban population and represent a variety of clinical settings for surveillance, general adult care, high-risk specialty services, and paediatric care (12).

**Figure 1.**
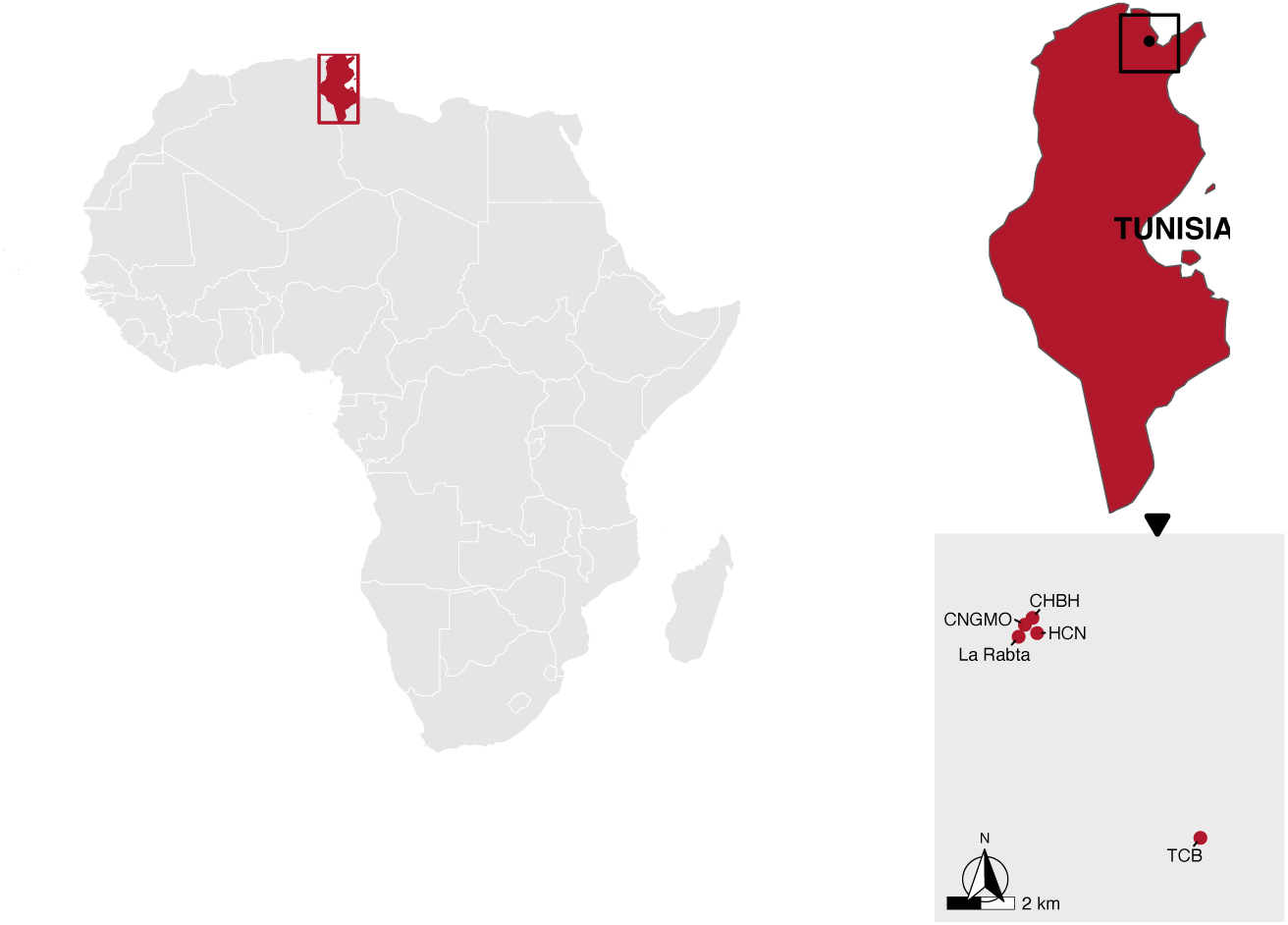
Location of the five tertiary hospitals participating in TARSS. Maps depict the geographic context and the hospital locations for TARSS sentinel network included in this study. Tunisia is highlighted within Africa. Tunisia, with the Greater Tunis study area and Greater Tunis showing the five hospitals (Charles Nicolle University Hospital [HCN], La Rabta Hospital [La Rabta], Children’s Hospital Bechir Hamza [CHBH], Centre National de Greffe de la Moelle Osseuse [CNGMO]) in Tunis and Trauma Centre and Burns [TCB] in Ben Arous. Basemap was sourced from Natural Earth (public domain). Points indicate the hospital locations.

### Data collection and inclusion criteria

In TARSS, bloodstream infection (BSI) surveillance captures febrile patients admitted to hospitals, where blood culture and AST are exclusively performed (i.e., these are not available locally outside hospital settings). Urinary tract infection (UTI) surveillance includes both symptomatic and asymptomatic hospital patients. While TARSS laboratories record both positive and negative cultures, only positive isolates with AST are reported to the NRL of AMR for national-level analysis and were thus included in this study.

The dataset included previously collected and anonymized isolate-level data from the five TARSS sites. Variables collected included: date of specimen collection, specimen type (blood, urine, wound and soft tissue, respiratory, CSF, stool, genital), referring hospital, patient demographics (age and sex), and AST profiles. To avoid duplication, only the first isolate per patient per specimen type per pathogen within the same year and hospital was included, in adherence with WHO GLASS de-duplication recommendations (13). The de-duplication process is centrally applied at the NRL during data cleaning and analysis, after all isolates were submitted to TARSS by the participating hospitals. We used the same de-duplication approach to ensure consistency with national surveillance data.

Isolates were cultured and identified using routine diagnostic methods in the hospital, and AST was performed in line with international standards (EUCAST). Data from 2014 to 2018 were retrospectively validated by the NRL of AMR after TARSS establishment in 2019. Validation was performed through data review and data quality checks, rather than retesting, to ensure consistency in reporting, completeness, and alignment with national surveillance criteria. Two distinct infection acquisition statuses were defined: community-acquired (CAI) infections, defined by the isolation of the pathogen from an outpatient or on days 0, 1, or 2 of current admission as an inpatient, or healthcare-associated infections (HAI), defined as an infection occurring >2 days since hospital admission.

Missing data, including unrecorded age or sex, were included in the overall analysis and missing data points were labeled as “not available” for transparency. However, these records were excluded from stratified and subgroup analyses relating to age and sex.

### Bacterial identification and antimicrobial susceptibility testing

Clinical specimens (e.g., blood, urine, respiratory, wound, CSF) were collected from hospitalized patients as part of routine diagnostic care at participating TARSS laboratories. Bacterial identification was performed using conventional microbiological techniques and API identification strips (bioMérieux). AST was conducted for each pathogen using disk diffusion methods following EUCAST guidelines. To ensure laboratories provide quality-assured standardized results, all laboratories participate in an external quality assessment scheme coordinated by the NRL of AMR and the NCC.

### AMR analysis

Disk diffusion data were analysed using the AMR package *v2.1.0* in R *v4.2.2* to interpret diffusion zone diameters according to EUCAST breakpoint tables 14.0 (14), and to calculate the proportion resistant for key pathogen-antibiotic combinations (15). We reported resistance, for each pathogen-antibiotic combination, as the proportion of isolates tested that were interpreted as resistant “R”. Susceptible “S” and susceptible increased exposure “I”, were reported separately, following EUCAST recommendations (16). The antibiotics analysed in the study were based on the WHO AWaRe classification (Access/Watch/Reserve) and WHO GLASS (see **Supplementary Table S2–S3**) (17,18). For each pathogen, we assessed the proportion of isolates that were tested for each antibiotic, and pattern of missingness by year and hospital (**Supplementary Table S3**). We aimed to include antibiotics prioritised in the WHO GLASS protocol, but we excluded pathogen-antibiotic combinations for which susceptibility results were available for less than 40% of all recorded isolates, to support unbiased temporal comparisons, or where EUCAST interpretive breakpoints were not applicable across the full range of specimen types (e.g., fosfomycin). Colistin was excluded because the AST was performed using disk diffusion, which is not recommended by EUCAST guidelines (14).

For the classification of multi-antibiotic resistance (MDR), we used the definition by Magiorakos *et al.,* which includes non-susceptibility (R+I) to ≥1 agent in ≥3 antimicrobial categories (19). Antibiotics that were intrinsically inactive for a given organism were excluded. For *P. aeruginosa*, where 97.1% of isolates meet the definition of MDR due to many drugs having only ‘I’ and ‘R’ categories and no ‘S’ interpretation possible using EUCAST breakpoints, MDR was defined as resistance (R) to ≥1 agent in ≥3 antimicrobial categories. To avoid misclassification from sparse AST, isolates were considered classifiable for MDR if AST results were available for ≥5 classes for *E. coli* and *K. pneumoniae*, and ≥4 classes for *P. aeruginosa* and *Acinetobacter* spp., and unclassifiable isolates were excluded.

We evaluated co-resistance within two groups: carbapenem-resistant (CR) isolates, defined as resistant to any carbapenem tested (imipenem/meropenem/ertapenem for *K. pneumoniae* and *E. coli*; imipenem/meropenem for *Acinetobacter* spp. and *P. aeruginosa*), and third-generation cephalosporin-resistant (3GCR), defined as resistant to ceftazidime or cefotaxime (for *K. pneumoniae* and *E. coli* only). For each pathogen, we calculated the proportion of isolates resistant to other antibiotics among CR and among 3GCR isolates, using the number of isolates tested for the respective antibiotic as the denominator.

For temporal trends in resistance, we plotted annual resistance proportions stratified by hospital ward and specimen type. Individual wards were summarised into groups: Intensive care unit (ICU), Emergency, Surgical, Medical, Paediatrics and neonates, while Specimen types were summarised into: Blood, Urine, Respiratory, Wound and soft tissues, and Other (including CSF, stool, genital tract). To assess linear trends over time, we applied the Cochran–Armitage trend test to analyse the resistance proportion for each specimen type or ward category, using *prop.trend.test* function in base R. Specimens with ≥10 isolates per year were plotted and <10 were censored. We also used the Cochran–Armitage trend test to assess linear trends in resistance over time for 3GC and carbapenem phenotypes, with ≥10 isolates per year.

Temporal changes in 3GCR and CR were assessed in more detail for the four pathogens. For each pathogen, we first identified CR isolates, defined as above, “R” to ≥1 carbapenem tested. We then constructed resistance profiles using a pathogen-specific panel of non-carbapenem antibiotics beyond β-lactams, including ciprofloxacin, trimethoprim-sulfamethoxazole, aminoglycosides (amikacin and gentamicin), tigecycline, and fosfomycin. Since not all isolates were tested against every antibiotic, we applied two rules to reduce bias from incomplete testing:

i. panel selection, an antibiotic was retained in the resistance profile panel only if recorded AST results (S/I/R) were available for ≥70% of CR isolates for that pathogen (see **Supplementary Table S4**); (ii) isolate completeness rule, an isolate was included in resistance profile analyses only if it has recorded AST results for ≥70% of drugs in the retained panel. This prevents isolates from being misclassified as having simpler profiles because some antibiotics were not tested. Isolates with no resistance (i.e., ‘S’ or ‘I’ to all antimicrobials in the resistance panel) were classified as “none”. For each year, we calculated the proportion for each resistance panel and plotted them as stacked area charts. To improve readability, we displayed individually the seven most frequent profiles for each pathogen, and collapsed the remaining into “Other”. The annual denominator (N) represents the number of CR isolates contributing to the resistance panel calculation for that pathogen and year. Colours were selected to reflect mechanistic content (blue ciprofloxacin resistance, red aminoglycoside resistance, purple resistance to both, green trimethoprim-sulfamethoxazole resistance).

Ages were recorded in years. Isolates associated with implausible age values >120 (n=7) were excluded from age group analyses. We pooled the observed ages across all four pathogens (*K. pneumoniae*, *E. coli*, *P. aeruginosa*, and *Acinetobacter* spp.) and divided these into quintiles, resulting in five age groups for reporting (<6 years, 6–<30 years, 30–<52 years, 52–<67 years, and ≥67 years). As infectious disease epidemiology can vary substantially in neonates and infants, we further divided the <6 age group into neonates (0–<28 days), infants (29–365 days), and children 1–<6 years, resulting in seven age groups in total.

### Statistical analysis

For each pathogen-antibiotic combination, we conducted multivariable logistic regression to estimate the adjusted odds ratios (aORs) with 95% confidence intervals (CIs). Models were fit independently using the *glm* function in base R for each outcome variable (i.e. R versus S/I, consistent with EUCAST (16) for individual drugs, and MDR vs non-MDR). Models were sequentially developed to explore the contribution of hospital and ward variation, see **Table 1** for model specifications and covariate definitions. **Model 1** included specimen type, hospital, and a simplified ward categorization (ICU, Emergency, Other). **Model 2** included an interaction term for hospital and ward category. We compared models with and without hospital–ward interaction using Akaike Information Criterion (AIC) to identify the best-fitting model. **Model 3** extended **Model 2** by including sex, and **Model 4** further included age group as an ordered categorical variable. The latter was treated as exploratory due to substantial missing age data (**Table 2**). Primary **Model 2** was also fitted to 3GC susceptible and resistant pathogens (*E. coli* and *K. pneumoniae*) and carbapenem susceptible and resistant pathogens (*E. coli*, *K. pneumoniae*, *Acinetobacter* spp., and *P. aeruginosa*).

**Table 1.** Summary of multivariable logistic regression models applied in analysis. Note: Nested comparisons used likelihood ratio test (LRTs) and difference Akaike Information Criterion (ΔAIC); nonnested contrasts relied on AIC.

| Model | Outcome | Formula | Description |
| --- | --- | --- | --- |
| **M1** | Resistant | ~ Year + Specimen + Hospital + Ward | Base model |
| **M1'** | MDR |  |  |
| **M2** | Resistant | ~ Year + Specimen + Hospital * Ward | Primary model: hospital*ward interaction term |
| **M2'** | MDR |  |  |
| **M3** | Resistant | ~ Year + Specimen + Hospital * Ward + Sex | Adds sex (reference: male)<br>Exploratory due to $\geq 24\%$ missing data for each pathogen (see Table 2) |
| **M3'** | MDR |  |  |
| **M4** | Resistant | ~ Year + Specimen + Hospital * Ward + Sex + Age_group | Adds age group (reference category: 30–<52 years)<br>Exploratory due to $\geq 40\%$ missing data for each pathogen (see Table 2) |
| **M4'** | MDR |  |  |

**Table 2.** Distribution of clinical isolates reported to TARSS, stratified by epidemiological and demographic variables across the five hospitals in Tunisia, 2014–2022. Values represent number of isolates and their percentages (%) relative to the total number of isolates for each pathogen. CNGMO = Centre National de Greffe de la Moelle Osseuse; TCB= Trauma Centre and Burns; CHBH= Children’s Hospital Bechir Hamza; HCN= Charles Nicolle University Hospital; ICU= Intensive Care Unit.

| Variable Level | *K. pneumoniae* | *E. coli* | *Acinetobacter* spp. | *P. aeruginosa* |
| --- | --- | --- | --- | --- |
| **Total Isolates** | 14,325 | 35,525 | 5,597 | 9,679 |
| ***Hospital*** |  |  |  |  |
| CHBH | 1830 (12.8%) | 8071 (22.7%) | 396 (7.1%) | 2380 (24.6%) |
| CNGMO | 160 (1.1%) | 150 (0.4%) | 38 (0.7%) | 183 (1.9%) |
| HCN | 4634 (32.3%) | 13057 (36.8%) | 1309 (23.4%) | 1706 (17.6%) |
| La Rabta | 5221 (36.4%) | 12108 (34.1%) | 1947 (34.8%) | 2767 (28.6%) |
| TCB | 2480 (17.3%) | 2139 (6%) | 1907 (34.1%) | 2643 (27.3%) |
| ***Specimen Type*** |  |  |  |  |
| Blood | 1,906 (13.3%) | 1033 (2.9%) | 1126 (20.1%) | 817 (8.4%) |
| Respiratory | 1,506 (10.5%) | 55 (0.2%) | 1641 (29.3%) | 2470 (25.5%) |
| Urine | 7,217 (50.4%) | 29317 (82.5%) | 609 (10.9%) | 1265 (13.1%) |
| Wound & soft tissues | 2,036 (14.2%) | 265 (0.7%) | 874 (15.6%) | 2216 (22.9%) |
| Other | 1,660 (11.6%) | 4855 (13.7%) | 1347 (24.1%) | 2911 (30.1%) |
| ***Year*** |  |  |  |  |
| 2014 | 1783 (12.4%) | 4226 (11.9%) | 718 (12.8%) | 953 (9.8%) |
| 2015 | 1671 (11.7%) | 4414 (12.4%) | 595 (10.6%) | 1091 (11.3%) |
| 2016 | 1773 (12.4%) | 5386 (15.2%) | 672 (12%) | 1276 (13.2%) |
| 2017 | 2139 (14.9%) | 5849 (16.5%) | 733 (13.1%) | 1472 (15.2%) |
| 2018 | 1984 (13.8%) | 5328 (15%) | 687 (12.3%) | 1252 (12.9%) |
| 2019 | 164 (1.1%) | 243 (0.7%) | 522 (9.3%) | 494 (5.1%) |
| 2020 | 1758 (12.3%) | 3812 (10.7%) | 678 (12.1%) | 1437 (14.8%) |
| 2021 | 1866 (13%) | 3880 (10.9%) | 600 (10.7%) | 999 (10.3%) |
| 2022 | 1187 (8.3%) | 2387 (6.7%) | 392 (7%) | 705 (7.3%) |
| ***Hospital Ward*** |  |  |  |  |
| Emergency | 957 (6.7%) | 6723 (18.9%) | 115 (2.1%) | 180 (1.8%) |
| Gynaecology | 343 (2.4%) | 1242 (3.5%) | 30 (0.5%) | 21 (0.2%) |
| ICU | 2823 (19.7%) | 818 (2.3%) | 2959 (52.9%) | 3517 (36.3%) |
| Medical | 5300 (37%) | 14688 (41.3%) | 1135 (20.3%) | 2852 (29.5%) |
| Paediatrics & neonates | 1843 (12.9%) | 4846 (13.6%) | 391 (7%) | 826 (8.5%) |
| Surgical | 2665 (18.6%) | 6168 (17.4%) | 878 (15.7%) | 2163 (22.3%) |
| Outpatient | 178 (1.2%) | 405 (1.1%) | 14 (0.2%) | 42 (0.4%) |
| Other | 17 (0.1%) | 85 (0.2%) | 8 (0.1%) | 59 (0.6%) |
| Not available | 199 (1.4%) | 550 (1.5%) | 67 (1%) | 19 (0.2%) |
| ***Infection Type*** |  |  |  |  |
| Community acquired | 1162 (8.1%) | 3492 (9.8%) | 86 (1.5%) | 690 (7.1%) |
| Healthcare associated | 1147 (8%) | 954 (2.7%) | 620 (11.1%) | 1070 (11.1%) |
| Not available | 12016 (83.9%) | 31079 (87.5%) | 4891 (87.4%) | 7919 (81.8%) |
| ***Sex*** |  |  |  |  |
| Male | 5539 (38.6%) | 8892 (25.0%) | 2623 (46.9%) | 4558 (47.1%) |
| Female | 4822 (33.7%) | 16648 (46.9%) | 1325 (23.7%) | 2765 (28.6%) |
| Not available | 3964 (27.7%) | 9985 (28.1%) | 1649 (29.5%) | 2356 (24.3%) |
| ***Age group*** |  |  |  |  |
| 0–28 days | 188 (1.3%) | 308 (0.9%) | 33 (0.6%) | 60 (0.6%) |
| 29–365 days | 234 (1.6%) | 830 (2.3%) | 24 (0.4%) | 135 (1.4%) |
| 1–<6 years | 148 (1%) | 1238 (3.5%) | 255 (4.6%) | 366 (3.8%) |
| 6–<30 years | 320 (2.2%) | 2940 (8.3%) | 108 (1.9%) | 590 (6.1%) |
| 30–<52 years | 622 (4.3%) | 3081 (8.7%) | 203 (3.6%) | 316 (3.3%) |
| 52–<67 years | 732 (5.1%) | 2641 (7.4%) | 260 (4.6%) | 393 (4.1%) |
| ≥67 years | 735 (5.1%) | 2824 (7.9%) | 224 (4.0%) | 308 (3.2%) |
| Not available | 11346 (79.2%) | 21663 (61.0%) | 4490 (80.2%) | 7511 (77.6%) |

Reference categories for were selected to maximize interpretability and precision. Specimen type used blood as the reference category since it represents the most clinically severe presentation, is available for all four pathogens, and avoids sparse counts from rarer sources. Hospital-level effects were evaluated using HCN as the reference site because of its median resistance profile and the large sample size. HCN also hosts the NRL of AMR, which contributed to consistent laboratory and reporting methods across the surveillance period. ICU ward served as a reference category reflecting the highest-risk and most consistently defined setting across hospitals. Year was included as a continuous covariate to capture temporal trends. Male sex was taken as the reference (largest stratum), and age used the middle quintile (30–<52 years) category as reference, to which paediatric and older age extremes could be compared.

Model performances were assessed using AIC and Area Under the Receiver Operating Characteristic Curve (AUC). Based on AUC thresholds, model discrimination was classified as excellent (>0.9), acceptable (0.8–0.9), fair (0.7–0.8), or poor (<0.7) (20). Multicollinearity was evaluated using variance inflation factors (VIF) using *car* (*v3.1-3)*. A VIF of >5 indicates moderate collinearity; models exceeding this threshold were flagged for interpretive caution. We used likelihood ratio tests (LRTs) to compare nested models and AIC for non-nested models. The model with the best fit (lowest AIC and highest AUC) was retained for primary interpretation.

Adjusted odds ratios were estimated from the model coefficients using the *tidy()* function in the *broom* package (*v1.0.7*). All analyses were conducted using R version *v4.2.2*. All model outputs are available in the GitHub repository via Zenodo (DOI: 10.5281/zenodo.18412695) (21).

## Results

### Epidemiological and demographic characteristics

Between 2014 and 2022, a total of 35,525 *E. coli*, 14,325 *K. pneumoniae*, 9,679 *P. aeruginosa*, and 5,597 *Acinetobacter* spp. isolates were reported to TARSS from the five participating hospitals (**Table 2**). The majority of isolates originated from HCN and La Rabta, accounting for 66% of total submissions across all pathogens. Annual isolate counts were stable from 2014 to 2018, followed by a decline in 2019 and 2020 reflecting incomplete capture during TARSS implementation period and disruptions of COVID-19 (see **Table 2**).

Specimen type distribution reflected pathogen-specific clinical presentations and diagnostic pathways (**Table 2**). *E. coli* and *K. pneumoniae* were predominantly isolated from urine specimens (82.5% and 50.4% of isolates per pathogen, respectively), with the remaining *K. pneumoniae* divided fairly evenly across other specimen types (13.3% blood, 10.5% respiratory, 14.2% wound/soft tissue, 11.6% other) and *E. coli* being rarely detected outside of urine (2.9% blood, 0.2% respiratory, 0.7% wound/soft tissue, 13.7% other). *Acinetobacter* spp. was most frequently found in respiratory samples (29.3%), as was *P. aeruginosa* (25.5%). Higher proportions of *Acinetobacter* spp. (20.1%) and *K. pneumoniae* (13.3%) were isolated from blood cultures than *P. aeruginosa* (8.4%) or *E. coli* (2.9%), consistent with clinical concern for invasive infection.

The proportion of isolates from ICU was highest for *Acinetobacter* spp. (52.9%), followed by *P. aeruginosa* (36.3%), and these proportions were significantly greater than those observed for *K. pneumoniae* (19.7%) and *E. coli* (2.3%) (p<0.001). Many isolates originated from medical wards (41.3% of *E. coli*, 37% of *K. pneumoniae* vs 20.3% and 29.5% for each of *Acinetobacter* spp. and *P. aeruginosa*). A high proportion of *E. coli* isolates (18.9%) were from specimens taken in emergency departments, mostly from urine (96.5%) or Other specimen type (2.5%); 6.7% of *K. pneumoniae* (84.5% urine, 4% respiratory) were isolated from emergency department patients, and only 2.1% of *Acinetobacter* spp. and 1.8% of *P. aeruginosa*.

Demographic data were incomplete, with age missing in over 60% of records, particularly before 2018. Sex was missing in 27% of records (mainly in *P. aeruginosa* and *Acinetobacter* spp. isolates submitted by La Rabta). However, among with age records available (20,116 isolates), *K. pneumoniae* was predominantly isolated from urine in most age groups, whilst in neonates (0–28 days), a high proportion of the isolates identified were from blood samples (35%) (**Supplementary Figure S2**). Similarly, *E. coli* was predominantly isolated from urine across most age groups (**Supplementary Figure S2**). For *E. coli* and *K. pneumoniae*, the proportion of UTI was higher amongst females, 87% and 56%, respectively. Amongst *Acinetobacter* spp., the breakdown of specimen types was similar across age groups and sexes. *P. aeruginosa* was primarily isolated from respiratory specimens in those <30 years, shifting to other, mixed, specimen types in older age groups (**Supplementary Figure S2**).

Data on the onset of infection (CAI vs HAI) was only available for 14% of cases (for 2021 and 2022 from four hospitals: HCN, La Rabta, CHBH, and CNGMO. Among isolates with a known onset category, most *E. coli* and *K. pneumoniae* infections were CAI (78% and 50%, respectively), most of which were from urine specimens consistent with CAI-UTI, and with the high fraction of emergency department isolates for both pathogens. Most *Acinetobacter* spp. (87.8%) and *P. aeruginosa* (60.8%) were HAI infections, mainly from respiratory specimens (*Acinetobacter* spp.: 46% vs *P. aeruginosa*: 45%) (**Supplementary Figure S2**), consistent with the very low fraction of isolates originating in ED

### Overall AMR profiles and temporal trends

The proportion resistant for each pathogen and antibiotic, and temporal trends assessed by fitting a multiple logistic regression model for each pathogen-antibiotic combination (model M2 in **Table 1**), is summarized in **Table 3**. *K. pneumoniae* demonstrated a high proportion MDR (45.5%), and high proportions resistant to individual β-lactams including third-generation cephalosporins (3GC) (cefotaxime 46.9%; ceftazidime 43.6%), the fourth-generation cephalosporin (4GC) cefepime (40.5%), and aztreonam (40.5) (**Table 3**). Resistance to ciprofloxacin (38.9%) and gentamicin (36.9%) was also common, with much lower proportions resistant to amikacin (18.9%) or carbapenems (ertapenem, 16.7% and imipenem, 7.9%). All drugs showed a statistically significant temporal trend, with gentamicin decreasing slightly and all others (and MDR) increasing. The overall effects were small for most drugs (aOR<1.1), with the exceptions of amikacin (aOR 1.18, 95% CI 1.16–1.20, p= 5.71×10^−64^) which rose from 16.4% in 2014 to 27.6% in 2022, and imipenem (aOR 1.24, [95% CI, 1.2–1.28], p=2.8×10^−44^) which rose from 3.9% in 2014 to 13% in 2022 (**Table 3**).

**Table 3.** Mean annual resistance prevalence by antibiotic, stratified by pathogen. Total indicates the number of isolates with AST reported for that antibiotic across the surveillance period. Column three gives the percentage resistant for each pathogen-antibiotic combination, calculated as the mean of annual resistance proportions across 2014–2022, with the range (minimum to maximum) of annual values in brackets. Temporal trend was assessed based on the effect for ‘year’ estimated from the primary model (interaction model M2 and M2’), presented as adjusted odds ratio (aOR), 95% CI and p-value. aOR>1 indicates an increasing trend in resistance; aOR<1 indicates a decreasing trend. Significant increasing trends, with aOR>1.1 and p<0.05, are highlighted.

| Pathogen and antibiotic | Total isolates tested | Annual percentage resistant, mean [range] | Temporal trend, aOR [95% CI] and p-value |
| --- | --- | --- | --- |
| *K. pneumoniae* |  |  |  |
| **Multidrug-resistant** | 12,594 | 45.5% [40.9–49] | 1.03 [1.02–1.05], p=9.66x10<sup>-05</sup> |
| **Beta-lactams** |  |  |  |
| Ceftazidime | 12,472 | 43.6 [34.6–49.9] | 1.09 [1.08–1.11], p=1.32x10<sup>-30</sup> |
| Cefotaxime | 11,646 | 47.0 [40.8–56.0] | 1.04 [1.03–1.06], p=4.9x10<sup>-08</sup> |
| Cefepime | 7,574 | 40.5 [33.1–46.2] | 1.04 [1.02–1.06], p=5.54x10<sup>-05</sup> |
| Ertapenem | 12,222 | 16.7 [11.4–25.6] | 1.03 [1.01–1.06], p=0.0028 |
| Imipenem | 12,052 | 7.90 [3.9–13.2] | **1.24 [1.20–1.28], p=2.84x10<sup>-44</sup>** |
| Aztreonam | 11,625 | 40.5 [34.4–44.5] | 1.02 [1.01–1.04], p=0.0003 |
| Piperacillin-tazobactam | 8,163 | 36.4 [25.9–46.9] | **1.14 [1.11–1.16], p=3.67x10<sup>-31</sup>** |
| **Aminoglycosides** |  |  |  |
| Amikacin | 12,811 | 18.9 [10.4–28.0] | **1.18 [1.16–1.21], p=5.71x10<sup>-64</sup>** |
| Gentamicin | 10,688 | 36.9 [32.9–43.3] | 0.97 [0.96–0.99], p=0.000741 |
| **Other** |  |  |  |
| Ciprofloxacin | 12,216 | 38.9 [33.3–44.8] | 1.05 [1.04–1.07], p=1.36x10<sup>-10</sup> |
| Trimethoprim-sulfamethoxazole | 11,479 | 42.1 [35.8–52.6] | 1.02 [1.01–1.04], p=0.001 |
| ***E. coli*** |  |  |  |
| **Multidrug-resistant** | 32,814 | 17.5% [12.7–20.4] | 0.98 [0.97–0.99], p=0.000957 |
| **Beta-lactams** |  |  |  |
| Ampicillin | 22,243 | 70.7 [68.2–72.9] | 0.97 [0.96–0.99], p=0.000101 |
| Ceftazidime | 27,849 | 12.9 [8.2–16.3] | 1.05 [1.04–1.07], p=5.68x10<sup>-11</sup> |
| Cefotaxime | 26,793 | 17.0 [11.5–19.9] | 0.97 [0.96–0.98], p=5.73x10<sup>-05</sup> |
| Cefepime | 19,451 | 15.0 [10.1–19.7] | 0.94 [0.93–0.96], p=2.41x10<sup>-11</sup> |
| Ertapenem | 31,675 | 0.6 [0.4–1.1] | **1.13 [1.07–1.20], p=4.02x10<sup>-05</sup>** |
| Imipenem | 32,323 | 0.2 [0.0–0.5] | **1.13 [1.01–1.27], p=0.0330** |
| Aztreonam | 29,022 | 13.1 [7.4–16.5] | 0.97 [0.96–0.98], p= 0.0006 |
| Piperacillin-tazobactam | 21,211 | 10.5 [4.3–14.4] | **1.14 [1.11–1.16], p= 8.42x10<sup>-38</sup>** |
| **Aminoglycosides** |  |  |  |
| Amikacin | 32,118 | 4.7 [1.4–11.6] | **1.30 [1.27–1.33], p=1.45x10<sup>-108</sup>** |
| Gentamicin | 25,280 | 14.2 [8.2–18.2] | 0.96 [0.94–0.97], p=1.79x10<sup>-08</sup> |
| **Other** |  |  |  |
| Ciprofloxacin | 30,794 | 24.2 [18.8–26.3] | 1.01 [1.00–1.02], p=0.0976 |
| Tigecycline | 23,680 | 1.2 [0.0–2.9] | **1.24 [1.18–1.30], p=2.09x10<sup>-17</sup>** |
| Trimethoprim-sulfamethoxazole | 29,303 | 37.4 [27.1–43.1] | 0.93 [0.92–0.94], p=4.52x10<sup>-41</sup> |
| ***Acinetobacter spp.*** |  |  |  |
| **Multidrug-resistant** | 2,624 | 85.1% [78.2–91.9] | 1.08 [1.02–1.13], p=0.0043 |
| **Beta-lactams** |  |  |  |
| Imipenem | 4,232 | 82.3 [73.0–91.1] | 1.09 [1.05–1.13], p=7.9x10<sup>-06</sup> |
| **Aminoglycosides** |  |  |  |
| Amikacin | 4,838 | 80.4 [76.4–85.7] | 1.04 [1.01–1.08], p=0.0085 |
| Gentamicin | 4,264 | 80.9 [74.2–85.3] | 1.07 [1.04–1.11], p=4.84x10<sup>-05</sup> |
| **Other** |  |  |  |
| Ciprofloxacin | 4,823 | 87.9 [83.5–91.2] | 1.05 [1.01–1.09], p=0.0262 |
| Trimethoprim-sulfamethoxazole | 3,867 | 71.2 [59.8–84.7] | **1.16 [1.12–1.20], p=6.41x10<sup>-18</sup>** |
| ***P. aeruginosa*** |  |  |  |
| **Multidrug-resistant** | 8,104 | 27.1% [16.3–40.5] | 1.02 [0.99–1.05], p=0.051 |
| **Beta-lactams** |  |  |  |
| Ceftazidime | 6,271 | 16.1 [0.0–36.8] | **1.18 [1.13–1.22], p=5.77x10<sup>-19</sup>** |
| Cefepime | 6,659 | 30.4 [20.9–45.6] | 0.99 [0.97–1.02], p=0.9062 |
| Imipenem | 8,167 | 30.5 [19.9–44.0] | 1.00 [0.98–1.02], p=0.95 |
| Meropenem | 5,739 | 23.3 [8.2–37.1] | 1.07 [1.04–1.11], p=1.07x10<sup>-06</sup> |
| Aztreonam | 7,153 | 7.0 [3.6–13.7] | **1.16 [1.11–1.21], p=8.22x10<sup>-12</sup>** |
| Piperacillin-tazobactam | 7,052 | 30.2 [16.2–42.5] | **1.10 [1.07–1.13], p=1.67 x10<sup>-12</sup>** |
| **Aminoglycosides** |  |  |  |
| Amikacin | 8,439 | 25.1 [15.2–33.5] | 1.01 [0.98–1.04], p=0.2498 |
| **Other** |  |  |  |
| Ciprofloxacin | 8,282 | 26.2 [15.4–42.8] | 1.01 [0.99–1.04], p=0.0982 |

*E. coli* showed a much lower proportion MDR (17.5%), and much lower proportions resistant to individual antibiotics, compared with *K. pneumoniae* (**Table 3**). The most common resistance phenotype observed amongst *E. coli* was to ampicillin (70.7%), which is an expected (intrinsic) resistance of *K. pneumoniae* and therefore not tested. Resistance proportions for other β-lactams were much lower in *E. coli*, including 3GC (cefotaxime 17%, ceftazidime 12.9%), 4GC (cefepime 15%) and piperacillin-tazobactam (10.5%), and carbapenem resistance was rare (imipenem 0.2% and ertapenem 0.6%). Resistance to non-β-lactam antibiotics, trimethoprim-sulfamethoxazole (37.4%) and ciprofloxacin (24.2%) were common, but amikacin resistance (4.7%) and tigecycline resistance was rare (1.2%). Besides ciprofloxacin, all other drugs showed a statistically significant trend. Concerningly, the drugs with the lowest proportions resistant (ertapenem, imipenem, piperacillin-tazobactam, amikacin and tigecycline, all ≤10.5%) were all significantly increasing, with aORs above 1.1 (see **Table 3**).

*Acinetobacter* spp. exhibited a very high proportion of MDR (85.1%), and high proportions resistant for each individual antibiotic tested (71.2 to 87.9%, see **Table 3**). Significant increasing trends were observed for MDR (78.2% to 87% in 2022) and for each individual antibiotic (see **Table 3**). A moderate proportion of *P. aeruginosa* isolates were MDR (27.1%), and this showed an increasing trend (from 21.4% in 2014 to 34% in 2022). Concerningly, the most common resistances were to the drugs that are most important clinically, namely imipenem (30.5%), cefepime (30.4%) and piperacillin-tazobactam (30.2%), although of these drugs only piperacillin-tazobactam showed an increasing trend (**Table 3**).

### Factors associated with resistance

We used the multivariable logistic regression models described in **Table 1** to explore the relationship between resistance and year, specimen type, and ward (adjusting for hospital). In nearly all cases, Model 2 (including hospital*ward as an interaction term) provided a better fit than Model 1 (without this interaction term) (see **Supplementary Tables S6–13**). The exceptions were ampicillin, imipenem, ertapenem, piperacillin-tazobactam, and tigecycline in *E.coli*, where including the interaction term did not improve fit (**Supplementary Table S8**), but model estimates were similar (**Supplementary Table S5**). Therefore, for consistency, Model 2 was considered the primary model for all pathogen-antibiotic combinations, and results for these primary models are summarized in **Figure 2** (effect sizes for hospital and the interaction terms in these models are found on GitHub) (21).

**Figure 2.**
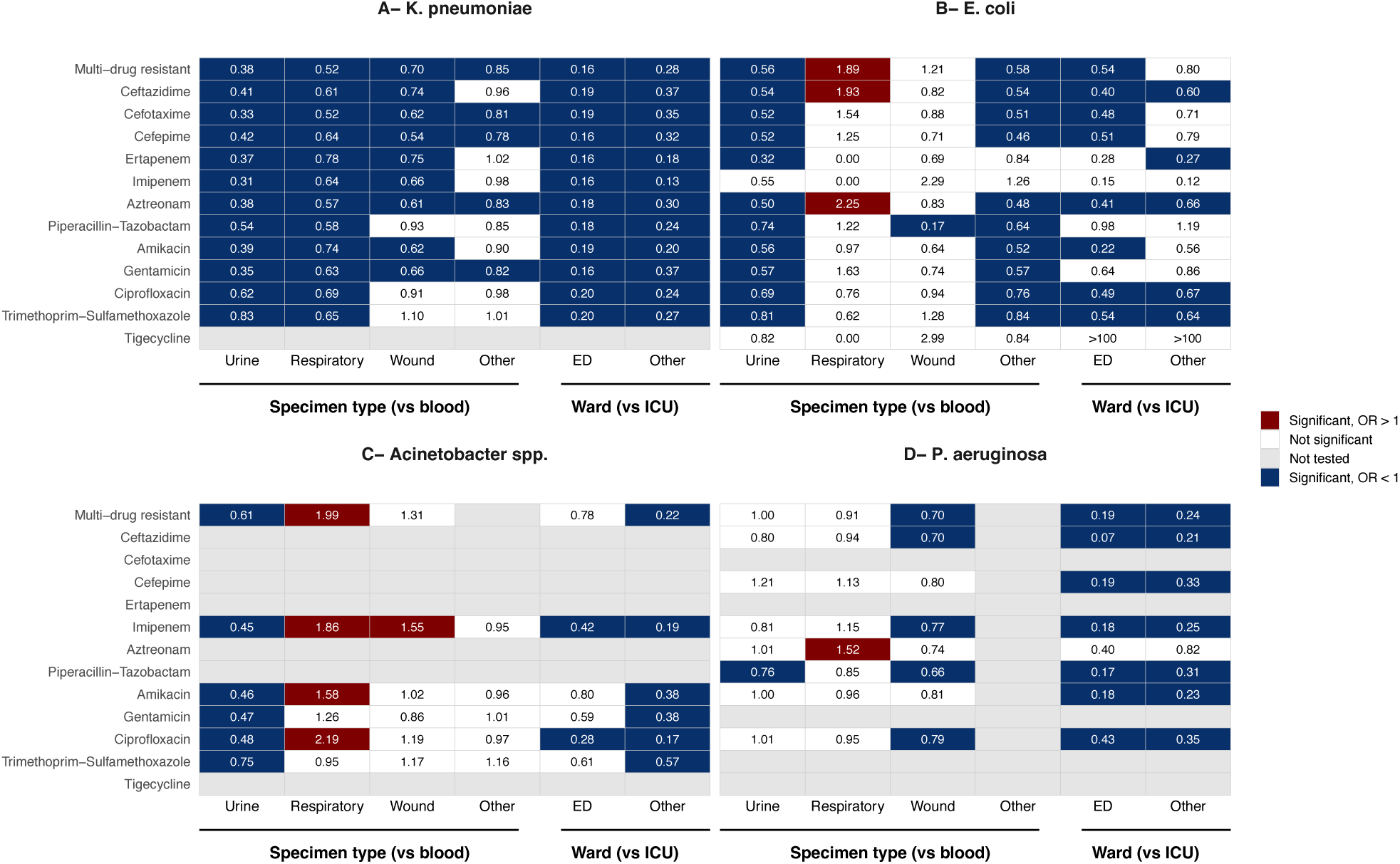
Adjusted effects from primary multivariable models (M2/M2’) for each pathogen-antibiotic combination. Each panel represents a pathogen. Rows indicate an individual antibiotic or multidrug resistance (as defined in Methods), and columns indicate model terms. Heatmap cells indicate adjusted odds ratios (aORs) for each term in Model 2 fit separately for each drug and pathogen; non-significant effects (p>0.05) are coloured white, significant effects (p<0.05) are coloured to indicate the directionality of the effect (red, aOR>1 or blue, aOR<1). Grey indicates the antibiotic was not modelled for this pathogen.

Overall, compared with ICU as the reference, emergency and other wards were significantly negatively associated with resistance across most antibiotic-pathogen combinations (**Figure 2**). Compared with blood as the reference specimen type, urine isolates were significantly negatively associated with resistance across most antibiotic-pathogen combinations (**Figure 2**). For *K. pneumoniae*, respiratory isolates were associated with lower odds of resistance to all drugs, compared with blood isolates. For the other pathogens, respiratory isolates were either not significantly different from blood isolates, or associated with higher odds of resistance compared with blood isolates; including aztreonam, ceftazidime, and MDR in *E. coli*, ciprofloxacin, amikacin, imipenem, and MDR in *Acinetobacter* spp., and aztreonam in *P. aeruginosa*. Wound or soft tissue isolates were associated with lower odds of resistance to all drugs for *K. pneumoniae* and most drugs for *P. aeruginosa*, but showed no association in *E. coli* and a significantly higher odds of resistance to imipenem amongst *Acinetobacter* spp.

As demographic data were incomplete, models including sex (Model 3) and age group (Model 4) were treated as exploratory only. Model 3 (fit to the 72.5% of samples with known sex) showed that, for *K. pneumoniae* and *E. coli*, females had a significantly lower odds of resistance to most drugs compared with males (**Supplementary Figure S3)**. Model 4 (fit to the 25% of samples with known age and sex) indicated significant differences in resistance by age group, with *K. pneumoniae* and *E. coli* isolated from young children having higher odds of resistance to most drugs, and *Acinetobacter* spp. and *P. aeruginosa* isolated from young children having lower odds, compared with the reference age group (30–<52 years, representing the middle quintile of age in years) (**Supplementary Figure S3).** For *E. coli,* older age groups (≥52 years) showed higher odds of resistance to most drugs and to MDR, however for the other pathogens older age groups were not generally associated with increased odds of resistance.

Temporal associations were discussed above and summarised in **Table 3**. These trends were consistent in directionality and scale across Models 1 through 3 **(Supplementary Table S5**). In the exploratory model M4 (fit to 25% of the data with age and sex data), some pathogen-antibiotic trends attenuated or changed direction; this is expected as the availability of age data varied over time, with earlier years (pre-2019 establishment of TARSS) lacking age data (**Supplementary Table S5**). To further explore temporal trends, we assessed annual resistance proportions for each pathogen and antibiotic at the ward level (**Figure 3**); these results are discussed below in separate sections for each pathogen.

**Figure 3.**
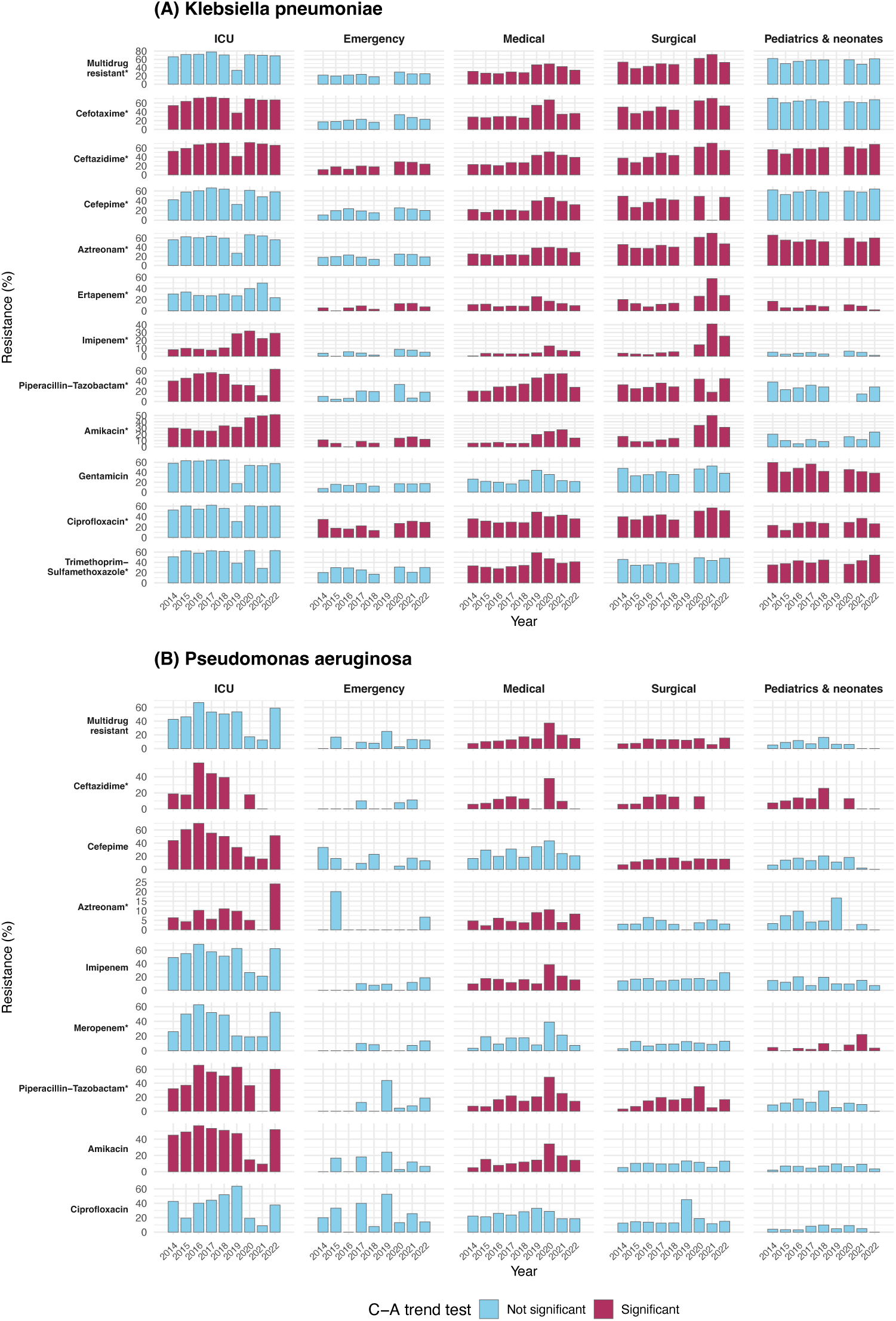
Ward-stratified AMR trends in antimicrobial resistance, 2014–2022. Resistance prevalence (%) is shown by year and ward (intensive care unit [ICU], emergency, medical, surgical, paediatrics and neonates) across (A) *K. pneumoniae* (B) *P. aeruginosa*. Significance of trends within wards was assessed with the Cochran–Armitage test of proportions (two-sided). Red color denotes significant trends, p<0.05; blue denotes not significant, p≥0.05. ‘*’ indicates a significant increasing overall temporal trend in the primary model (aOR > 1 and p< 0.05).

## Pathogen-specific findings

### Klebsiella pneumoniae

As noted above, increasing trends were detected for MDR and all individual antibiotics, except for gentamicin, which showed a weakly declining trend (**Table 3**). **Figure 3A** shows the temporal trends within wards, revealing that the greatest increases in proportions resistant occurred amongst isolates from the medical and surgical wards, with consistently high proportions amongst ICU and paediatric isolates. For example, in the ICU, the proportion MDR remained high and stable (ICU: 66% in 2014 to 68% in 2022; Cochran–Armitage trend test: χ^2^=0.2, p=0.6), whereas in medical wards, the proportion MDR increased from 31% in 2014 to 34% in 2022 (medical: χ^2^=69, p=6.7×10^−17^), and fluctuating in surgical wards from 38% to 72%. The same patterns were evident for ceftazidime, cefotaxime, cefepime, aztreonam, and ciprofloxacin (**Figure 3A**). Drugs with lower resistance proportions in ICU showed significant increases amongst ICU isolates over time, in particular imipenem resistance increased from 8% to 29% (ꭓ^2^= 99, p=8.99×10^−15^), ertapenem fluctuated from 23% to 49%, and amikacin from 29% to 51% (ꭓ^2^= 79, p=4.5×10^−19^). These drugs also showed increasing resistance amongst isolates from the medical and surgical wards, and had the strongest overall temporal trends in the M2 regression models (**Table 3**); increasing trends were also evident in all specimen types (**Figure 2**; **Supplementary Figure S5A)**. For these and other drugs (ceftazidime, ciprofloxacin), low but slowly increasing resistance was observed amongst isolates from patients in the emergency department, likely reflecting low but increasing resistance prevalence in the community. The overall decline in gentamicin resistance identified in the M2 model was not associated with any particular ward (**Figure 3A**), instead this trend was associated with specimen type, with resistant proportions remaining low and stable amongst urine isolates (mean 38.2%) compared with higher but steadily declining rates amongst blood, respiratory and wound/soft tissue isolates (**Supplementary Figure S5A**).

In total, 48% of *K. pneumoniae* isolates were 3GCR, rising slightly from 48.5% in 2014 to 51% in 2022. Amongst 3GCR *K. pneumoniae*, for every non-3GC antibiotic, the proportion resistant was significantly higher than the proportion resistant amongst 3GC susceptible (3GCS) strains (**Figure 4**). This includes ertapenem (32.0% amongst 3GCR vs 1.9% amongst 3GCS) and imipenem (15.3% vs 0.3%), as well as the non-beta-lactam antibiotics amikacin (34.3% vs 2.8%), gentamicin (70.6% vs 5.8%), ciprofloxacin (67.0% vs 11.3%) and trimethoprim-sulfamethoxazole (67.0% vs 15.8%). Most antibiotics showed an increasing trend amongst 3GCR strains but not amongst 3GCS (**Table 4**). Notably, resistance to ertapenem, imipenem and amikacin increased significantly after 2018 (**Figure 4**). There was also evidence of accumulation of resistance to multiple additional classes over time, with the proportion of 3GCR isolates co-resistant to ciprofloxacin, gentamicin, amikacin, and trimethoprim-sulfamethoxazole rising from 8% in 2014 to 32.4% in 2022. The proportion of 3GCR isolates co-resistant to all these antibiotics plus imipenem rose from 0.4% to 20% (**Supplementary Figure S6**).

**Figure 4.**
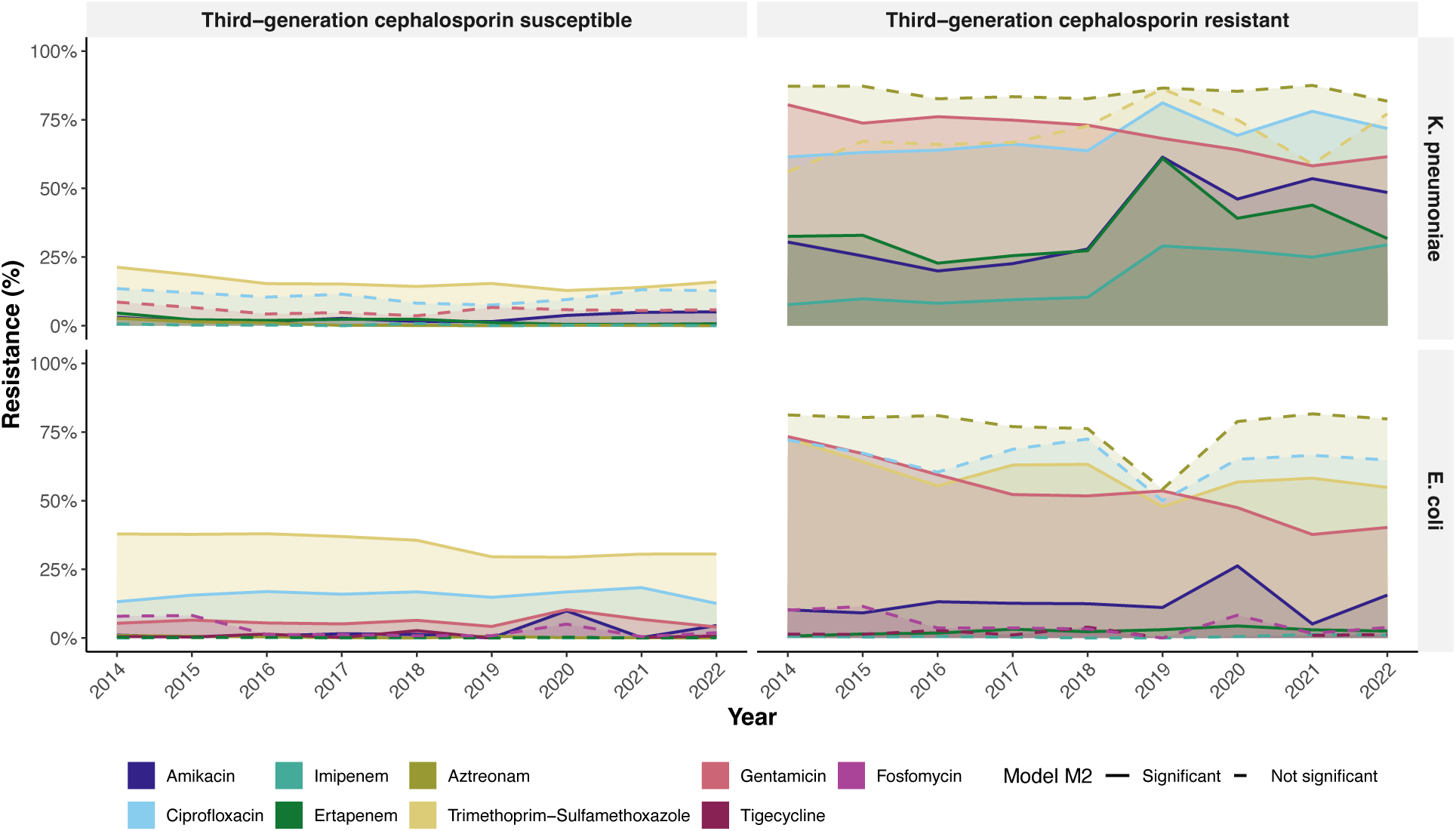
Resistance trends of third-generation cephalosporin phenotypes, 2014–2022. Third-generation cephalosporin phenotype (susceptible vs resistant). Coloured lines/shaded areas represent the annual percentage resistance for each antimicrobial. P-values are derived from Model M2. Line types indicate year effect in Model 2 (solid p <0.05; dashed p ≥0.05).

**Figure 5.**
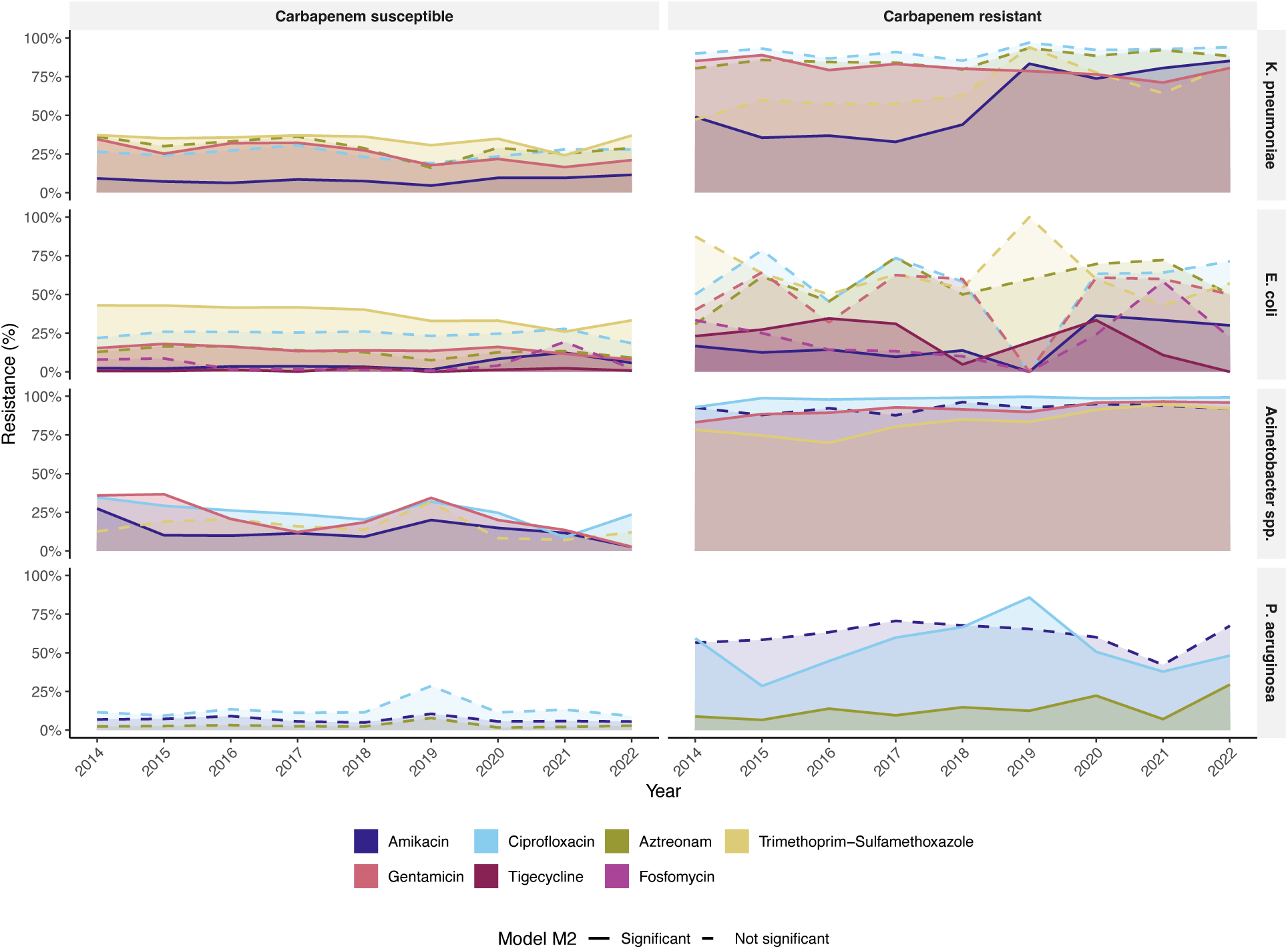
Resistance trends for carbapenem phenotypes, 2014–2022. Carbapenem phenotype (susceptible vs resistant). Coloured lines/shaded areas represent the annual percentage resistance for each antimicrobial. P-values are derived from Model M2. Line types indicate year effect in Model 2 (solid p <0.05; dashed p ≥0.05).

**Table 4.** Resistance and co-resistance profiles by third-generation cephalosporin and carbapenem susceptibility phenotype, stratified by pathogen. Third-generation cephalosporin resistance (3GCR) was defined as resistance to ≥1 to cefotaxime/ceftazidime. Carbapenem-resistant (CR) was defined as resistance to ≥1 carbapenem tested. For each phenotype group, percentage resistance (%R) was calculated as the number of isolates resistant over number of isolates with AST reported for that antibiotic. Differences in resistance between phenotype groups (3GCS vs 3GCR and CS vs CR) were assessed using a two-sample test of proportions where applicable; all comparisons were statistically significant (p ≤ 0.05). The odds ratio (aOR), 95% confidence interval, and p-value for the year effect were estimated by re-fitting the primary model M2 (hospital*ward) separately to 3GCR/ 3GCS and CR/CS isolates.

| Panel A. Third generation cephalosporin susceptible vs resistant |  |  |  |  |
| --- | --- | --- | --- | --- |
| Pathogen and antibiotic | 3GCS |  | 3GCR |  |
|  | %R | Temporal trends | %R | Temporal trends |
| *K. pneumoniae* |  |  |  |  |
| Beta-lactams |  |  |  |  |
| Cefepime | 0.80% | 0.76 [0.67–0.86],<br>p=1.96$\times 10^{-5}$ | 86.40% | 0.96 [0.92–1.01], p=0.101 |
| Ertapenem | 1.90% | 0.77 [0.70–0.84],<br>p=3.5$\times 10^{-9}$ | 32.00% | 1.06 [1.03–1.08],<br>p=3.6$\times 10^{-5}$ |
| Imipenem | 0.30% | 0.84 [0.68–1.04],<br>p=0.109 | 15.30% | 1.26 [1.22–1.30],<br>p=6.02$\times 10^{-4}$ |
| Aztreonam | 0.40% | 0.54 [0.43–0.67],<br>p=1.9$\times 10^{-8}$ | 84.00% | 1.00 [0.97–1.03], p=0.816 |
| Piperacillin-tazobactam | 11.3% | 1.07 [1.03–1.12],<br>p=0.00058 | 61.7% | 1.18 [1.14–1.23],<br>p=4.5$\times 10^{-22}$ |
| Meropenem | 1.20% | – | 22.50% | – |
| Aminoglycosides |  |  |  |  |
| Amikacin | 2.80% | 1.17 [1.10–1.25],<br>p=2.8$\times 10^{-7}$ | 34.30% | 1.20 [1.17–1.22],<br>p=1.84$\times 10^{-48}$ |
| Gentamicin | 5.80% | 0.98 [0.94–1.03],<br>p=0.414 | 70.60% | 0.89 [0.87–0.91],<br>p=2.47$\times 10^{-20}$ |
| Other |  |  |  |  |
| Ciprofloxacin | 11.30% | 1.00 [0.97–1.04],<br>p=0.916 | 67.00% | 1.04 [1.01–1.06],<br>p=0.0028 |
| Trimethoprim-sulfamethoxazole | 15.80% | 0.96 [0.93–0.99],<br>p=0.0049 | 67.00% | 1.02 [1.00–1.05], p=0.108 |
| *E. coli* |  |  |  |  |
| Beta-lactams |  |  |  |  |
| Ampicillin | 65.30% | 0.98 [0.97–1.00],<br>p=0.0099 | 99.40% | 1.23 [0.92–1.64], p=0.169 |
| Cefepime | 0.60% | 0.76 [0.70–0.84],<br>p=4.6$\times 10^{-9}$ | 89.20% | 0.87 [0.82–0.93],<br>p=6.2$\times 10^{-6}$ |
| Ertapenem | 0.20% | 1.03 [0.90–1.18],<br>p=0.68 | 2.40% | 1.18 [1.08–1.29],<br>p=2.3$\times 10^{-4}$ |
| Imipenem | 0.04% | 0.72 [0.52–1.00],<br>p=0.052 | 0.50% | 1.11 [0.92–1.35], p=0.262 |
| Aztreonam | 0.20% | 0.58 [0.49–0.69],<br>p=4.5$\times 10^{-10}$ | 77.30% | 0.98 [0.95–1.01], p=0.189 |
| Piperacillin-tazobactam | 7.50% | 1.16 [1.13–1.19],<br>p=5.1$\times 10^{-26}$ | 27.50% | 1.18 [1.14–1.23],<br>p=9.5$\times 10^{-18}$ |
| Aminoglycosides |  |  |  |  |

| **Amikacin** | 2.20% | **1.42 [1.36–1.49],**<br>**p=9.8×10<sup>-55</sup>** | 12.80% | **1.11 [1.07–1.16],**<br>**p=2.7×10<sup>-7</sup>** |
| --- | --- | --- | --- | --- |
| **Gentamicin** | 5.70% | 1.04 [1.01–1.06],<br>p=0.0056 | 56.00% | **0.85 [0.82–0.87],**<br>**p=3.2×10<sup>-28</sup>** |
| **Other** |  |  |  |  |
| **Ciprofloxacin** | 15.10% | 1.03 [1.01–1.04],<br>p=0.0022 | 66.30% | 0.98 [0.95–1.02], p=0.302 |
| **Tigecycline** | 1.10% | **1.45 [1.32–1.60],**<br>**p=3.0×10<sup>-14</sup>** | 2.10% | 1.09 [0.96–1.23], p=0.167 |
| **Trimethoprim-sulfamethoxazole** | 35.90% | **0.96 [0.95–0.97],**<br>**p=9.9×10<sup>-11</sup>** | 60.70% | **0.93 [0.90–0.96],**<br>**p=2.1×10<sup>-6</sup>** |
| **Fosfomycin** | 3.00% | – | 5.90% | – |
| **Panel B. Carbapenem susceptible vs resistant** |  |  |  |  |
| Pathogen and antibiotic | CS |  | CR |  |
|  | %R | Temporal trends | %R | Temporal trends |
| ***K. pneumoniae*** |  |  |  |  |
| **Beta-lactams** |  |  |  |  |
| **Cefotaxime** | 36.20% | 1.00 [0.99–1.02],<br>p=0.695 | 90.80% | 1.18 [1.10–1.27],<br>p=7.1×10<sup>-6</sup> |
| **Ceftazidime** | 32.50% | **1.06 [1.04–1.08],**<br>**p=4.2×10<sup>-10</sup>** | 90.90% | **1.32 [1.23–1.42],**<br>**p=2.4×10<sup>-14</sup>** |
| **Cefepime** | 31.90% | 1.00 [0.98–1.02],<br>p=0.943 | 88.60% | **1.23 [1.11–1.35],**<br>**p=4.1×10<sup>-5</sup>** |
| **Piperacillin-tazobactam** | 26.1% | **1.1 [1.07–1.13],**<br>**p=5.9x10<sup>-12</sup>** | 96.9% | 1.15 [0.9–1.4], p=0.24 |
| **Aminoglycosides** |  |  |  |  |
| **Amikacin** | 8.48% | **1.08 [1.05–1.12],**<br>**p=1.2×10<sup>-7</sup>** | 56.50% | **1.33 [1.28–1.39],**<br>**p=3.4×10<sup>-40</sup>** |
| **Gentamicin** | 27.40% | **0.94 [0.92–0.96],**<br>**p=4.9×10<sup>-10</sup>** | 80.40% | **0.91 [0.86–0.95],**<br>**p=1.63x10<sup>-4</sup>** |
| **Other** |  |  |  |  |
| **Ciprofloxacin** | 26.30% | 1.01 [0.99–1.03],<br>p=0.159 | 90.90% | 1.02 [0.95–1.09], p=0.576 |
| **Trimethoprim-sulfamethoxazole** | 34.60% | **0.97 [0.96–0.99],**<br>**p=0.0065** | 61.6% | 0.913 [0.784–1.063],<br>p=0.24 |
| ***E. coli*** |  |  |  |  |
| **Beta-lactams** |  |  |  |  |
| **Cefotaxime** | 17.10% | **0.96 [0.95–0.98],**<br>**p=4.6×10<sup>-6</sup>** | 64.90% | 1.27 [1.02–1.58], p=0.036 |
| **Ceftazidime** | 12.80% | **1.05 [1.03–1.07],**<br>**p=1.1×10<sup>-9</sup>** | 65.80% | **1.34 [1.08–1.66],**<br>**p=0.0085** |
| **Cefepime** | 15.30% | **0.94 [0.92–0.96],**<br>**p=7.7×10<sup>-13</sup>** | 58.70% | 1.03 [0.81–1.31], p=0.797 |
| **Piperacillin-tazobactam** | 10.70% | **1.14 [1.11–1.16],**<br>**p=8.4×10<sup>-34</sup>** | 63.90% | **1.36 [1.02–1.83], p=0.037** |
| **Aminoglycosides** |  |  |  |  |
| **Amikacin** | 4.72% | **1.29 [1.26–1.33],**<br>**p=7.0×10<sup>-99</sup>** | 21.40% | **1.54 [1.26–1.88],**<br>**p=2.3×10<sup>-5</sup>** |
| **Gentamicin** | 14.60% | 0.95 [0.94–0.97],<br>p=1.0×10<sup>-9</sup> | 52.30% | 1.14 [0.95–1.38], p=0.16 |
| **Ciprofloxacin** | 24.90% | 1.01 [0.99–1.02],<br>p=0.303 | 62.00% | 1.06 [0.91–1.25], p=0.438 |
| **Other** |  |  |  |  |
| **Tigecycline** | 1.24% | **1.28 [1.22–1.35],**<br>**p=2.2×10<sup>-20</sup>** | 21.10% | 0.86 [0.69–1.07], p=0.168 |
| **Trimethoprim-sulfamethoxazole** | 38.80% | **0.93 [0.92–0.94],**<br>**p=2.4×10<sup>-40</sup>** | 56.20% | 0.91 [0.78–1.06], p=0.243 |
| **Fosfomycin** | 5.00% | – | 25.60% | – |
| ***Acinetobacter* spp.** |  |  |  |  |
| **Aminoglycosides** |  |  |  |  |
| **Amikacin** | 15.00% | **0.89 [0.80–0.98],**<br>**p=0.016** | 92.50% | 1.03 [0.97–1.09], p=0.329 |
| **Gentamicin** | 24.30% | **0.87 [0.80–0.95],**<br>**p=0.0013** | 91.50% | **1.22 [1.15–1.29],**<br>**p=1.1×10<sup>-11</sup>** |
| **Other** |  |  |  |  |
| **Ciprofloxacin** | 26.00% | **0.90 [0.83–0.98],**<br>**p=0.019** | 98.20% | **1.36 [1.20–1.55],**<br>**p=1.3×10<sup>-6</sup>** |
| **Trimethoprim-sulfamethoxazole** | 17.20% | 1.09 [0.97–1.21],<br>p=0.135 | 82.00% | **1.19 [1.13–1.25],**<br>**p=6.2×10<sup>-11</sup>** |
| ***P. aeruginosa*** |  |  |  |  |
| **Beta-lactams** |  |  |  |  |
| **Ceftazidime** | 7.54% | **1.11 [1.04–1.18],**<br>**p=0.0010** | 53.50% | **1.37 [1.29–1.46],**<br>**p=3.9×10<sup>-24</sup>** |
| **Cefepime** | 10.90% | 0.99 [0.95–1.03],<br>p=0.551 | 70.70% | 1.00 [0.95–1.05], p=0.969 |
| **Piperacillin-tazobactam** | 9.39% | **1.14 [1.08–1.19],**<br>**p=1.3×10<sup>-7</sup>** | 70.30% | **1.20 [1.15–1.27],**<br>**p=4.2×10<sup>-13</sup>** |
| **Aztreonam** | 2.61% | 1.04 [0.96–1.13],<br>p=0.352 | 13.50% | **1.24 [1.17–1.31],**<br>**p=2.4×10<sup>-14</sup>** |
| **Aminoglycosides** |  |  |  |  |
| **Amikacin** | 6.61% | 0.98 [0.93–1.03],<br>p=0.401 | 62.80% | 1.00 [0.96–1.05], p=0.874 |
| **Other** |  |  |  |  |
| **Ciprofloxacin** | 12.40% | 1.00 [0.97–1.04],<br>p=0.941 | 51.40% | **1.05 [1.01–1.09],**<br>**p=0.0128** |

Overall, 15.3% of *K. pneumoniae* isolates were carbapenem-resistant. CR remained stable over time at 15.4% in 2014 and 15.1% in 2022. CR *K. pneumoniae* showed much higher resistance than carbapenem-susceptible (CS) isolates, with >80% resistance observed for all drugs tested, except amikacin, which was 56.5% resistant amongst CR isolates and 8.5% resistant amongst CS isolates (**Table 4**). Resistance to multiple additional drug classes appears to be accumulating over time, with co-resistance of CR isolates to the combination of amikacin, gentamicin, ciprofloxacin, and trimethoprim-sulfamethoxazole increasing from 12% in 2014 to 64.5% in 2022 (**Supplementary Figure S7**).

### Escherichia coli

As noted above, increasing trends were observed for ertapenem, imipenem, amikacin, piperacillin-tazobactam, and tigecycline (**Table 4**). For imipenem and ertapenem, the increase in resistance was most evident amongst ICU isolates (from 0% to 1.7%, and 1.2% to 3%, respectively) (**Supplementary Figure S4A**), and resistance remained rare in non-ICU isolates (≤2.4%). For amikacin and piperacillin-tazobactam, the annual proportion resistant was sustained at high levels amongst ICU isolates (mean 12.8% and 17.7% per year, respectively) and the trend was driven by increases in non-ICU wards (e.g. from 0% to 4.4%, and from 8% to 12%, in emergency department isolates) (see **Supplementary Figure S4A**) and observed across blood and urine isolates (**Supplementary Figure S5**).

In total, 17.5% of *E. coli* isolates were 3GCR, which remained stable over the period, fluctuating between 12% and 20%. Among 3GCR *E. coli*, all antibiotics showed a significantly higher proportion resistant compared with 3GCS *E. coli*, with very high resistance to ciprofloxacin (66.3% amongst 3GCR vs 15.1% amongst 3GCS), trimethoprim-sulfamethoxazole (60.7%) (**Table 4**, **Figure 4A**). Amikacin and piperacillin-tazobactam showed an increasing trend amongst both 3GCS and 3GCR isolates (**Figure 4**, **Table 4**), reaching 15.6% amikacin resistant and 33% piperacillin-tazobactam resistant in 2022. Carbapenem resistance remained rare, but increasing, amongst 3GCR isolates (**Table 4**), reaching 2.5% ertapenem resistant and 1.1% imipenem resistant in 2022. Most 3GCR *E. coli* remained susceptible to fosfomycin (6%) and tigecycline (2%), with no significant increase amongst 3GCR or 3GCS isolates (**Table 4**).

Whilst carbapenem resistance was rare in *E. coli* (**Table 3**), CR *E. coli* showed >50% co-resistance to most other antibiotics, except for fosfomycin (25.6%), amikacin (21.4%) and tigecycline (21%). Combined resistance to at least two of amikacin, ciprofloxacin, and trimethoprim-sulfamethoxazole increased from 42.9% to 71.4%; meanwhile, the majority of CR *E. coli* isolates remained resistant to at least one of these agents (**Supplementary Figure S7**).

### Acinetobacter *spp*

As noted above, very high proportions (>70%), and increasing trends, were detected for MDR and all individual antibiotics (**Table 3**). The highest rates, with modest increases, were amongst ICU isolates, whilst medical wards showed intermediate but increasing rates (**Supplementary Figure S4B**). For example, amongst ICU isolates MDR increased from 85% in 2014 to 94% in 2022 (Cochran–Armitage trend test: ꭓ^2^=8, p=0.003). By 2022, resistance prevalence in ICU isolates was high for multiple antibiotics, imipenem (92.5%), ciprofloxacin (94%), amikacin (89%), and gentamicin (92%) (all p<0.001). Amongst isolates from medical wards, MDR increased from 68% to 81% in (ꭓ^2^=31, p=2.1×10^−8^), with parallel increases for several antibiotics (gentamicin: 54% to 60%; ꭓ^2^=14; p=0.00012, ciprofloxacin: 75% to 76%; ꭓ^2^=30, p=3.8×10^−8^, and amikacin: 59% to 61%; ꭓ^2^=48, p=3.48×10^−12^). These trends were also associated with specimen type: isolates from respiratory specimens showed a high and increasing trend in MDR (76% to 91%; χ²=14.7, p=0.0001), and across all antibiotics, exceeding 80% by 2022 (p<0.001) (**Supplementary Figure S5)**. Isolates from paediatric wards showed the lowest proportions resistant, consistent with lower odds of resistance amongst children (**Supplementary Figure S3)**, although the proportions resistant were still concerningly high (MDR: 75%, imipenem: 76.4%, gentamicin 76.4%, ciprofloxacin 76.4%).

Most *Acinetobacter* spp. isolates were CR (82.3%), and amongst these, co-resistance was very high (>80%) for ciprofloxacin, amikacin, gentamicin, and trimethoprim-sulfamethoxazole (**Table 4**). The proportions resistant to other antibiotics were much lower among CS isolates (15%-26%; see **Table 4**), highlighting the strong correlation of resistance determinants across multiple drug classes within the *Acinetobacter* population. Concerningly, the proportion of CR *Acinetobacter* spp. also resistant to amikacin, gentamicin and ciprofloxacin increased from 77% in 2014 to 91% in 2022 (**Supplementary Figure S7**).

### Pseudomonas aeruginosa

In *P. aeruginosa,* most antibiotics showed resistance proportions in the range 16–30%, with moderate overall increasing trends (**Table 3**), and significantly higher odds of resistance amongst ICU isolates (**Figure 2**). Although ICU isolates were overall more resistant, amongst ICU isolates the proportions resistant declined over time for several drugs, and the overall increasing trends were associated more with medical and surgical wards (**Figure 3B**). Wound and soft tissue specimens had lower odds of resistance for several antibiotics (**Figure 2**), but with resistance to all antibiotics increasing among such specimens (**Supplementary Figure S5**). For example, MDR increased from 7% to 44% (ꭓ^2^=81.5, p=1.73×10^−19^), piperacillin-tazobactam from 5% to 44% (ꭓ^2^=124, p=8.3×10^−29^) and imipenem from 12% to 46% (ꭓ^2^=40.6, p=1.8×10^−10^) (**Supplementary Figure S5**).

Among CR *P. aeruginosa* isolates, co-resistance was substantially high to several anti-pseudomonal agents (70% piperacillin-tazobactam, 70.7% cefepime, 62.8% amikacin, 53% ceftazidime, 51.4% ciprofloxacin), but much lower for aztreonam (13%) (**Table 4**). Combined resistance to amikacin and ciprofloxacin was sustained at high levels (mean 91.8%) amongst *P. aeruginosa* (**Supplementary Figure S7**).

## Discussion

Here, we present a nine-year, multi-hospital analysis of AMR trends across five tertiary TARSS hospitals, to augment the sparse surveillance data currently available from African countries (22,23). By analysing >65,000 de-duplicated isolates with ward, specimen, and hospital data, we elucidated temporal, spatial, and clinical variation in resistance with a granularity rarely reported in this region.

We show that resistance increased for many pathogen-antibiotic combinations between 2014 and 2022 (**Table 3**). The odds of MDR increased over time for *K. pneumoniae* (aOR 1.03), *Acinetobacter* spp. (aOR 1.08), and *P. aeruginosa* (aOR 1.02), but decreased in *E. coli* (aOR 0.98) (**Table 3**). Isolates from ICU and invasive specimens (blood, respiratory) had higher odds of AMR, whereas urine was associated with the lowest odds of AMR (**Figure 2**).

Relative to regional comparators, our findings are directionally concordant. GLASS data for the WHO Eastern Mediterranean Region (EMR) report the highest prevalence of carbapenem resistance amongst bloodstream *Acinetobacter* spp. (70.3%) and the lowest in *E.coli* (4.6%) (18,24). Similarly, a Tunisian bloodstream infection study reported rising 3GC resistance in Enterobacterales and high prevalence of carbapenem resistance amongst *Acinetobacter* spp. (25). We extend this evidence by analysing all clinical specimen types and quantifying associations with factors relating to clinical setting. In our dataset, *K. pneumoniae* showed a sustained increase in the proportions resistant to 3GC (reaching 50% in 2022) and carbapenem resistance (reaching 13% for imipenem in 2022), with a higher odds of resistance for blood isolates and those collected from patients in ICU (**Figure 3A**). In this context, amongst 3GCR isolates resistance to several other commonly tested antibiotics was frequent, and amongst CR isolates resistance to most tested antibiotics was >80%, with the exception being amikacin (56.5% resistant) (**Table 4**). This high frequency of co-resistance indicates a limited range of antibiotics with retained *in vitro* activity, which severely constrains empiric treatment choices. We also observed a concerning upward trend for amikacin resistance in the ICU and several other wards (**Figure 3A**), which is concerning. *E. coli,* on the other hand, displayed a slight decline in 3GC resistance (cefotaxime: 15% to 12% by 2022) and several first-line antibiotics, while carbapenem resistance remained rare (<1%) (**Supplementary Figure S4A**). However, the odds of resistance were higher in blood specimens and those from ICU patients, with proportions exceeding 20% for most drugs tested (**Supplementary Figure S4A; Supplementary Figure S5**). While GLASS data reveal alarming levels of 3GCR *E. coli* in the Eastern Mediterranean (44.8%) and particularly in Africa (70%) (26,27). The low levels of resistance may, in part, be due to Tunisia’s ongoing stewardship efforts; as of 2024, they maintained a 60% Access-group antibiotic target from 2000 to 2018, reflecting prudent antibiotic use and compliance with their national essential medicines list (27). Among non-fermenters, *Acinetobacter* spp. exhibited a high increase in resistance across most antibiotics (>81% by 2022), especially in ICU and respiratory specimens **(Supplementary Figure S4B; Supplementary Figure S5).** *Acinetobacter* spp. is problematic in the EMR with the highest reported figures globally (66.5%) and rising 11.3% annually (28). Our findings are consistent with prior surveillance and clinical studies reported from Tunisia, which also observed high multidrug resistance exceeding 80% while revealing some susceptibility to trimethoprim-sulfamethoxazole in *Acinetobacter* spp. (29). Additionally, *P. aeruginosa* showed an upward trend in resistance to anti-pseudomonal antibiotics (aztreonam, piperacillin-tazobactam), with blood and ICU as the highest-risk settings (**Figure 3B and Supplementary Figure S5**).

In *P. aeruginosa*, where “I” is common and ICU/blood isolates are tested against multiple classes, the Magiorakos MDR definition (R +I) (19) tends towards saturation, resulting in total (100%) MDR. Accordingly, we modified the 2012 MDR definition to R only (β1 agent in β3 antimicrobial classes), aligning it to our single-antibiotic outcomes (%R) reporting as recommended in recent EUCAST guidelines (19,30). Magiorakos MDR definition is the most common criterion used in hospitals and the surveillance networks, such as the European Centre for Disease Prevention and Control (31). However, surveillance programmes in Germany and Switzerland have issued their own MDR definitions primarily in the aim of guiding IPC measures (32,33). The German nationwide definition focuses on four key classes for severe infections (anti-pseudomonal penicillins, ESBL cephalosporins, carbapenems, and quinolones) and grades MDR into categories, which then uses this data to apply isolation precautions. While no national consensus guidelines exist in Switzerland, the University Hospital Zurich developed a definition in 2008 that classifies MDR as resistance to three of five categories (including aminoglycosides). Wolfensberger *et al.* (31) have shown that differences in MDR definitions affect MDR classification, and they advocate that definitions should take into consideration local resistance rates and epidemiological priorities. We also suggest that, for interpretation and comparison, surveillance systems may be better served by focusing on single antibiotic %R and co-resistance patterns, and standardising R-only MDR definitions, following recent EUCAST recommendations (16).

Isolate submission dipped in 2019–2020 with partial recovery in 2021 and 2022. This decline coincided with the COVID-19 service reprioritisation. Major networks such as the European Antimicrobial Resistance Surveillance Network documented perturbing routine AMR surveillance and test volumes during this period (34–36). We therefore interpret 2019–2020 cautiously and modelled year as a continuous covariate, to summarise average temporal changes across the period.

The concentration of resistance in ICU and blood/respiratory specimens is consistent with the settings where invasive devices use such as ventilators and antibiotic exposure as key risk factors. Accordingly, our findings support existing recommendations that IPC priorities in these settings include device stewardship (tight control of ventilator, central-line, and catheter device-days), environmental controls (with wet controls *for P. aeruginosa* and intensified dry-surface decontamination for *Acinetobacter* spp.), unit-level antibiograms by ward, targeted audits, and outbreak readiness (37–40). Beyond the expected ICU pattern, our adjusted model revealed hospital-ward heterogeneity (**Figure 2**). The interaction effects reinforce the usefulness of ward-specific antibiograms and empirical guidance.

Our analysis showed the existence of marked differences in susceptibility depending on where pathogens were isolated. While the results were exploratory, given the large amount of missing data on sex and age, female sex was associated with significantly lower odds of resistance and MDR across *K. pneumoniae* and *E. coli,* as well as resistance to ciprofloxacin in *P. aeruginosa* (**Supplementary Figure S3**). Several studies concur with these findings and argue that certain behavioural factors may place men at more risk of resistance (41–43). It was also striking in the *E. coli* model, female sex had lower odds of resistance. A plausible factor is that women are prone to CA or HA urinary tract infections from the microflora, which are more likely to be susceptible, while such infections are rare in men and when they occur, are typically HAI and more likely to be caused by hospital strains, which are more resistant (44). Moreover, age effects showed higher odds of resistance in neonates, infants, and children for *K. pneumoniae*, and mixed effects for *E. coli* (higher in infants and older adults) (**Supplementary Figure S3**). A similar study from China reported higher rates of carbapenem-resistant Enterobacterales in the neonatal group, potentially due to lower immunity and a greater propensity to bacterial infections (45). *Acinetobacter* spp. and *P. aeruginosa* in older age groups are at elevated risk of resistance, as they are more likely to undergo invasive procedures in healthcare settings and have chronic conditions, which increase their risk of colonisation and infection (46,47). These patterns are consistent with Waterlow *et al.*’s large-scale European analysis, which demonstrates that resistance prevalence varies by age and sex, with the most apparent effect in *E. coli* and *K. pneumoniae* (48). While their study focuses on bloodstream infections, our cohort spans all specimen types, which accentuates the specimen mix influence. These findings underscore the importance of demographically and clinically stratified stewardship and surveillance.

## Strengths and Limitations

Key strengths of this study include its longitudinal, multicentre design, which spans nine years (2014–2022), and the participation of five sentinel hospitals in TARSS. Our large sample size enhances precision, allows robust comparisons within tertiary-care hospitals in Tunis and Ben Arous, and captures a wide range of clinical and ward-level variations. We applied multivariable logistic regression models that included hospital and ward level and their interaction, thereby reducing confounding from institutional differences. Additionally, we applied GLASS de-duplication and leveraged TARSS standardisation and EQA process, which improved comparability of microbiological data across participating laboratories.

However, several limitations must be acknowledged. First, demographic data completeness was suboptimal: age was absent in over 60% of records prior to 2018, and infection type (community versus healthcare-associated) was only available for 14% of records and only from 2021 onwards. Second, our unit of analysis was isolates, deduplicated to represent one isolate per specimen type and patient per hospital and year, which may be skewed by localised transmission clusters within hospitals, or by movement of patients between hospitals. Third, the study is based on routine passive laboratory surveillance, where ascertainment of isolates is influenced by diagnostic practices and testing availability. Therefore, our estimations are limited to the proportion resistant amongst cultured isolates, and not the incidence of resistant infections in the patient population. Fourth, phenotypic resistance was not linked to resistance mechanisms; as a result, we could not assess the contribution of genetic determinants or clonal lineage to the observed resistance trends. Fifth, we lacked data on antibiotic usage at the patient and facility levels, which precludes exploring the association between resistance rates and antimicrobial exposure. Finally, as with all hospital-based surveillance, community-acquired infections are underrepresented. Notably, isolates originating from patients in the emergency department, which presumably include the highest fraction of community-acquired infections, had much lower proportions of resistance.

## Conclusion

This nine-year multi-hospital analysis provides, to our knowledge, the most comprehensive description of AMR patterns in Tunisian tertiary care hospitals. *K. pneumoniae* showed sustained increase in 3GC and carbapenem resistance, *Acinetobacter* spp. remained highly resistant across all clinically relevant classes, and *P. aeruginosa* exhibited lower but rising resistance to key anti-pseudomonal antibiotics. *E. coli* showed high low and stable rates of 3GC resistance, whereas carbapenem resistance was rare but rising. Across pathogens, the resistance burden was the highest in ICUs and invasive specimens, highlighting the importance of ward-level antibiograms and stratified stewardship guidelines to guide empirical therapy as well as targeted IPC measures in high-risk settings. Taken together, these findings underscore the importance of sustained harmonised surveillance to inform national AMR policy.

## Data Availability

The data underlying this study are derived from the Tunisian Antimicrobial Resistance Surveillance System (TARSS) and are not publicly available due to data governance and confidentiality restrictions. Aggregated data supporting the findings are presented in the manuscript and supplementary materials. Model outputs and analysis code are publicly available on GitHub (doi/10.5281/zenodo.18412695).

## Acknowledgements

We thank our collaborators from the Tunisian hospitals from CHBH, La Rabta, CNGMO, and TCB, and the NRL of AMR for their invaluable contributions, especially for providing the data. This work was conducted as part of TARSS under the supervision of the NRL of AMR. We further acknowledge the sustained support of the NRL and the MOH in Tunisia for their leadership and continued commitment to AMR surveillance activities.

## Author contributions

Conceptualization, DI, KEH; methodology, DI, LTP, KEH; data provision, IB, MZ, HS, LT, WA; data collation and extraction, IB, SK; formal analysis, DI; visualization, DI; writing—original draft preparation, DI; writing—review and editing, DI, KEH, LTP, EFN, IB, HS, MZ, LT, WA; supervision, KEH. All authors have read and approved the final version of the manuscript.

## References

1. O’Neill J. Tackling drug-resistant infections globally: final report and recommendations [Internet]. Government of the United Kingdom; 2016 May [cited 2022 Aug 4]. Available from: https://apo.org.au/node/63983

2. WHO [Internet]. World Health Organization; [cited 2021 Apr 29]. WHO | Global action plan on AMR. Available from: http://www.who.int/antimicrobial-resistance/global-action-plan/en/

3. WHO [Internet]. World Health Organization; [cited 2020 Sep 9]. GLASS | Global Antimicrobial Resistance Surveillance System (GLASS). Available from: http://www.who.int/glass/en/

4. Aenishaenslin C, Häsler B, Ravel A, Parmley J, Stärk K, Buckeridge D. Evidence needed for antimicrobial resistance surveillance systems. Bulletin of the World Health Organization. 2019 Apr 1;97(4):283–9.

5. Iskandar K, Molinier L, Hallit S, Sartelli M, Hardcastle TC, Haque M, et al. Surveillance of antimicrobial resistance in low- and middle-income countries: a scattered picture. Antimicrob Resist Infect Control. 2021 Mar 31;10:63.

6. Data for Lower middle income, Tunisia | Data [Internet]. [cited 2025 Jul 15]. Available from: https://data.worldbank.org/?locations=XN-TN

7. Tunisia: National action plan to fight against antimicrobial resistance (French) [Internet]. [cited 2023 Jan 23]. Available from: https://www.who.int/publications/m/item/tunisia-national-action-plan-to-fight-against-antimicrobial-resistance

8. WHO bacterial priority pathogens list, 2024: Bacterial pathogens of public health importance to guide research, development and strategies to prevent and control antimicrobial resistance [Internet]. [cited 2024 Nov 20]. Available from: https://www.who.int/publications/i/item/9789240093461

9. Dziri O, Dziri R, Ali El Salabi A, Chouchani C. Carbapenemase Producing Gram-Negative Bacteria in Tunisia: History of Thirteen Years of Challenge. Infect Drug Resist. 2020 Nov 23;13:4177–91.

10. Antimicrobial resistance TrACSS Tunisia 2022 country profile [Internet]. [cited 2025 Mar 3]. Available from: https://www.who.int/publications/m/item/Antimicrobial-resistance-tracss-tun-2022-country-profile

11. STROBE [Internet]. [cited 2025 Jul 2]. STROBE. Available from: https://www.strobe-statement.org/

12. Statistics | INS [Internet]. [cited 2025 Dec 18]. Available from: https://www.ins.tn/statistiques/111

13. GLASS guide to preparing aggregated antimicrobial resistance data files [Internet]. [cited 2025 Nov 7]. Available from: https://www.who.int/publications/i/item/WHO-DGO-AMR-2016.6

14. EUCAST: Clinical breakpoints and dosing of antibiotics [Internet]. [cited 2021 Sep 14]. Available from: https://eucast.org/clinical_breakpoints/

15. Antimicrobial Resistance Data Analysis [Internet]. [cited 2025 Jul 28]. Available from: https://amr-for-r.org/

16. eucast: S, I and R and surveillance of AMR [Internet]. [cited 2025 Oct 5]. Available from: https://www.eucast.org/eucast_news/news_singleview?tx_ttnews%5Btt_news%5D=599&cHash=946cd7aca91faf703fcd4ef2099e82a3

17. AWaRe classification of antibiotics for evaluation and monitoring of use, 2023 [Internet]. [cited 2025 Mar 3]. Available from: https://www.who.int/publications/i/item/WHO-MHP-HPS-EML-2023.04

18. WHO [Internet]. World Health Organization; [cited 2020 Nov 4]. GLASS | Global antimicrobial resistance and use surveillance system (GLASS) report. Available from: http://www.who.int/glass/resources/publications/early-implementation-report-2020/en/

19. Magiorakos AP, Srinivasan A, Carey RB, Carmeli Y, Falagas ME, Giske CG, et al. Multidrug-resistant, extensively drug-resistant and pandrug-resistant bacteria: an international expert proposal for interim standard definitions for acquired resistance. Clinical Microbiology and Infection. 2012 Mar 1;18(3):268–81.

20. Çorbacıoğlu ŞK, Aksel G. Receiver operating characteristic curve analysis in diagnostic accuracy studies: A guide to interpreting the area under the curve value. Turk J Emerg Med. 2023 Oct 3;23(4):195–8.

21. Itani D. danaitani/Model_outputs: TARSS AMR model outputs v1.0.0 [Internet]. Zenodo; 2026 [cited 2026 Feb 16]. Available from: https://zenodo.org/doi/10.5281/zenodo.18412695

22. Tadesse BT, Ashley EA, Ongarello S, Havumaki J, Wijegoonewardena M, González IJ, et al. Antimicrobial resistance in Africa: a systematic review. BMC Infect Dis. 2017 Sep 11;17(1):616.

23. Tornimbene B, Eremin S, Abednego R, Abualas EO, Boutiba I, Egwuenu A, et al. Global Antimicrobial Resistance and Use Surveillance System on the African continent: Early implementation 2017–2019. African Journal of Laboratory Medicine. 11(1):1594.

24. Talaat M, Zayed B, Tolba S, Abdou E, Gomaa M, Itani D, et al. Increasing Antimicrobial Resistance in World Health Organization Eastern Mediterranean Region, 2017–2019. Emerg Infect Dis. 2022 Apr;28(4):717–24.

25. Kanzari L, Ferjani S, Meftah K, Zribi M, Mezghani S, Ferjani A, et al. Bacterial Pathogens and Antibiotic Resistance in Bloodstream Infections in Tunisia: A 13-Year Trend Analysis. Tropical Medicine and Infectious Disease. 2025 Jun;10(6):164.

26. Global antibiotic resistance surveillance report 2025: summary [Internet]. [cited 2026 Feb 16]. Available from: https://www.who.int/publications/i/item/B09585

27. World Health Organization Regional Office for the Eastern Mediterranean. Prevention and control of antimicrobial resistance in the Eastern Mediterranean Region – a progress report, 2024 [Internet]. World Health Organization Regional Office for the Eastern Mediterranean; Available from: https://applications.emro.who.int/docs/9789292743666-eng.pdf

28. WHO EMRO - World Antimicrobial Resistance Awareness Week 2025: Act Now, to protect our present and secure our future [Internet]. [cited 2026 Jan 13]. Available from: https://www.emro.who.int/media/news/world-antimicrobial-resistance-awareness-week-2025-act-now-to-protect-our-present-and-secure-our-future.html

29. Nadia J, Wejdene M, Remy A. B, Meriam G, Cherifa C, Rachida G, et al. Temporal Variation in Antibiotic Resistance of Acinetobacter baumannii in a Teaching Hospital in Tunisia: Correlation with Antimicrobial Consumption. [cited 2026 Jan 13]; Available from: https://openmicrobiologyjournal.com/VOLUME/13/PAGE/106/FULLTEXT/

30. eucast: S, I and R and surveillance of AMR [Internet]. [cited 2025 Jul 3]. Available from: https://www.eucast.org/eucast_news/news_singleview?cHash=946cd7aca91faf703fcd4ef2099e82a3&tx_ttnews%5Btt_news%5D=599&utm_source=chatgpt.com

31. Wolfensberger A, Kuster SP, Marchesi M, Zbinden R, Hombach M. The effect of varying multidrug-resistence (MDR) definitions on rates of MDR gram-negative rods. Antimicrob Resist Infect Control. 2019 Nov 28;8(1):193.

32. RKI - Homepage - [Internet]. [cited 2026 Jan 13]. Available from: https://www.rki.de/DE/Themen/Infektionskrankheiten/infektionskrankheiten-node.html?blob=publicationFile;

33. Tissot F, Widmer AF, Kuster SP, Zanetti G. Enterobacteriaceae mit Breitspektrum Beta-Laktamasen (ESBL) im Spital: Neue Empfehlungen Swissnoso 2014.

34. Farfour E, Clichet V, Péan de Ponfilly G, Carbonnelle E, Vasse M. Impact of COVID-19 pandemic on blood culture practices and bacteremia epidemiology. Diagn Microbiol Infect Dis. 2023 Sep;107(1):116002.

35. Ansari S, Hays JP, Kemp A, Okechukwu R, Murugaiyan J, Ekwanzala MD, et al. The potential impact of the COVID-19 pandemic on global antimicrobial and biocide resistance: an AMR Insights global perspective. JAC Antimicrob Resist [Internet]. 2021 Apr 8 [cited 2021 Sep 13];3(2). Available from: https://academic.oup.com/jacamr/article/3/2/dlab038/6217452

36. Antimicrobial resistance in the EU/EEA (EARS-Net) - Annual epidemiological report for 2021 [Internet]. 2022 [cited 2025 Sep 23]. Available from: https://www.ecdc.europa.eu/en/publications-data/surveillance-antimicrobial-resistance-europe-2021

37. Global report on infection prevention and control 2024 [Internet]. [cited 2025 Sep 8]. Available from: https://www.who.int/publications/i/item/9789240103986

38. CDC. Infection Control. 2024 [cited 2025 Sep 23]. Disinfection and Sterilization Guideline. Available from: https://www.cdc.gov/infection-control/hcp/disinfection-and-sterilization/index.html

39. CDC. Healthcare-Associated Infections (HAIs). 2024 [cited 2025 Sep 23]. Considerations for Reducing Risk: Water in Healthcare Facilities. Available from: https://www.cdc.gov/healthcare-associated-infections/php/toolkit/water-management.html

40. SHEA. Compendium of Strategies to Prevent Healthcare-Associated Infections in Acute Care Hospitals – SHEA [Internet]. [cited 2025 Sep 23]. Available from: https://shea-online.org/compendium-of-strategies-to-prevent-healthcare-associated-infections-in-acute-care-hospitals/

41. Sahuquillo-Arce JM, Selva M, Perpiñán H, Gobernado M, Armero C, López-Quílez A, et al. Antimicrobial Resistance in More than 100,000 Escherichia coli Isolates According to Culture Site and Patient Age, Gender, and Location. Antimicrobial Agents and Chemotherapy [Internet]. 2011 Jan 10 [cited 2025 Dec 26]; Available from: https://journals.asm.org/doi/10.1128/aac.00765-10

42. Kodde C, Bonsignore M, Köhler J, Schwegmann K, Nachtigall I. Males are at higher risk of colonization and infection with multi-drug-resistant organisms than females. Journal of Hospital Infection. 2025 Jan 1;155:88–94.

43. Batheja D, Goel S, Charani E. Understanding gender inequities in antimicrobial resistance: role of biology, behaviour and gender norms. BMJ Glob Health [Internet]. 2025 Jan 20 [cited 2025 Dec 26];10(1). Available from: https://gh.bmj.com/content/10/1/e016711

44. Livermore DM, Nichols T, Lamagni TL, Potz N, Reynolds R, Duckworth G. Ciprofloxacin-resistant Escherichia coli from bacteraemias in England; increasingly prevalent and mostly from men. J Antimicrob Chemother. 2003 Dec 1;52(6):1040–2.

45. Fu P, Xu H, Jing C, Deng J, Wang H, Hua C, et al. Bacterial Epidemiology and Antimicrobial Resistance Profiles in Children Reported by the ISPED Program in China, 2016 to 2020. Microbiology Spectrum. 2021 Nov 3;9(3):e00283–21.

46. National trends in hospital, long-term care and outpatient Acinetobacter baumannii resistance rates | Microbiology Society [Internet]. [cited 2025 Dec 26]. Available from: https://www.microbiologyresearch.org/content/journal/jmm/10.1099/jmm.0.001473

47. Theodorakis N, Feretzakis G, Hitas C, Kreouzi M, Kalantzi S, Spyridaki A, et al. Antibiotic Resistance in the Elderly: Mechanisms, Risk Factors, and Solutions. Microorganisms. 2024 Oct;12(10):1978.

48. Waterlow NR, Cooper BS, Robotham JV, Knight GM. Antimicrobial resistance prevalence in bloodstream infection in 29 European countries by age and sex: An observational study. PLOS Medicine. 2024 Mar 14;21(3):e1004301.

